# A Comparative Analysis of Area-Based Socioeconomic Measures: Implications for Future Equity-focused Public Health Response

**DOI:** 10.1101/2025.05.12.25327460

**Authors:** Cesar Aviles-Guaman, Ada T. Kwan, Priya B. Shete

## Abstract

Effectively identifying communities in need of public health resources is critical for addressing health disparities. However, clear strategies for doing so and prioritizing resources are not well established. As area-based socioeconomic measures (ABSMs), which include indices that capture determinants of health for a specific geographical unit, increasingly gain traction for guiding policy and resource allocation, it is essential to understand how different ABSMs perform in relation to health outcomes of interest. This proof-of-concept study illustrates an approach to compare how different ABSMs as place-based indicators are associated with disease outcomes. Using monthly COVID-19 public health surveillance data for 2020-2021 at the census tract level in California, we qualitatively and quantitatively compare five prominent ABSMs: California Healthy Places Index, Area Deprivation Index, Social Vulnerability Index, Index of Concentration at the Extremes, and Home Owners’ Loan Corporation (HOLC) “redlining” grades. Our findings demonstrate that no single ABSM consistently aligned with COVID-19 case and mortality rates across geographies or time, highlighting the importance of selecting measures based on context, data availability, data quality, and the specific health outcome of interest. Moreover, our analysis revealed that associations between poor health outcomes and proxy measures for historical disinvestment and racial discrimination suggest these patterns are important to identify when developing equitable public health strategies. This work underscores the potential for public health decision-makers and implementers to use both qualitative and quantitative approaches to select among ABSMs for targeting interventions more effectively.

## Introduction

A challenge in public health is how to identify populations to effectively address health disparities.^1,2^ In the United States (US), health disparities are a priority public health concern and have been most commonly described using a single factor such as race, ethnicity, age, or income.^3–7^ Yet health-related disparities are multifaceted, influenced by a complex interplay of broader factors often referred to as *social determinants of health* (SDOH). Defined by the World Health Organization as “the conditions in which people are born, grow, live, work and age, [including] the forces and systems shaping the conditions of daily life”,^8^ SDOH have further been shown to have persisting, intergenerational effects on the wellbeing of historically marginalized populations and racial/ethnic minority groups.^9^ While traditional approaches to addressing health disparities have often relied on demographic and socioeconomic factors, they present several limitations for effectively targeting health-related interventions. In addition to high rates of missingness for demographic and socioeconomic variables in individual-level surveillance data, these factors create challenges in implementing interventions solely based on individual or household-level categories, do not account for the contributing roles of environmental factors, and are often presented without the intersectional dimensions of stratification.^5,9,10^ Moreover, they often lack the nuance needed for priority setting in a comprehensive manner within public health.

In response to these limitations, area-based socioeconomic measures (ABSMs) have emerged as promising tools for capturing single or multiple dimensions of community health. ABSMs also include composite, place-based indices that typically aggregate various social, economic, structural, and anthropogenic indicators at different geographic levels, such as a neighborhood or census tract. In Great Britain, New Zealand, and several other high-income countries, ABSMs are commonly used as organizing principles for health-related interventions.^11^ In contrast to individual or household characteristics, ABSMs represent a paradigm shift in how we conceptualize and quantify health-related disparities, which can be useful for allocating resources during needs-based public health activities. However, many ABSMs exist, and a growing number continue to be constructed, each with its own specific design, set of indicators, methodologies, and geographic level of data availability. Reconciliation of these differences and similarities is necessary to effectively leverage the use of ABSMs in public health.

Several comparative analyses of ABSMs exist.^12–18^ In a recent systematic scoping review of commonly used ABSMs in the US since 2015, Trinidad et al. identified 15 distinct tools with notable differences in how these indices define the socioeconomic spectrum of advantage to disadvantage.^12^ Some examples include: the Area Deprivation Index (ADI) which captures social risk factors and has been widely utilized in resource allocation to identify areas with relative levels of neighborhood deprivation and vulnerability^19^; the Social Vulnerability Index (SVI) sourced from the Centers for Disease Control and Prevention’s Agency for Toxic Substances and Disease Registry (CDC/ATSDR), which has been used extensively in the US to prioritize resources in disaster planning, preparedness, and response^20^; the Index of the Concentration at the Extremes (ICE) developed by scholars at Harvard University, which evaluates income levels and focuses on the extremes within a population.^21^ While many of these ABSMs at their core include similar data such as from the American Community Survey (ACS), they each differ in their exact composition of constituents, method of validation, and geographic level in which data is available.^12^ Still other ABSMs incorporate historical data to explore the enduring impact of structural disadvantage on public health outcomes.^22^ One example is the Home Owners’ Loan Corporation (HOLC) “redlining” grades from the 1930s, which are colored maps that have been widely used as a proxy for historic disinvestment and racial discrimination.^19,22,23^ While the HOLC maps did not directly drive discriminatory mortgage lending practices by the HOLC, as more recent research has shown that many of these loans were already issued before the creation of the maps, the maps have been shown to reflect prevailing racial and economic biases of that time.^23,24^ Various studies detail further the relationship between HOLC grades and shaping demographic and spatial patterns, as well as racial, socioeconomic, and health disparities that have persisted to present day in American cities.^25,26^

An increasing number of studies have examined the relationships between ABSMs and health outcomes.^10,27,28^ Health disparities central to the COVID-19 pandemic prompted researchers and policymakers to leverage ABSMs to better understand and respond to disparities in COVID-19 outcomes.^29,30^ Since the onset of the pandemic, certain populations were disproportionately affected by COVID-19 infection, hospitalization, death, and vaccine disparities.^31–37^ In an analysis across 3,125 US counties and county equivalents, Tipirneni et al. (2022) found positive associations between COVID-19 incidence and mortality rates with four different ABSMs: the SVI, the ADI, the COVID-19 Community Vulnerability Index (CCVI), and the Minority-Health-Social Vulnerability Index (MH-SVI).^16^ Because of limited access to health data at more granular levels, only a few studies have examined ABSMs with health outcomes at a higher data resolution than county level. In a cohort study of 19,495 veterans living in California, Wong et al. (2023) found that the California Healthy Places Index (HPI) was a significant predictor of COVID-19 hospitalization risk among veterans.^38^ Specifically, Black and non-Hispanic white veterans living in lower HPI (less opportunity for living a healthy life) neighborhoods experienced higher hospitalization rates. For Black veterans, this association persisted even after adjusting for Black segregation; while for Hispanic veterans, lower HPI was not significantly associated with hospitalization after accounting for Hispanic segregation. These findings suggest that the HPI, like the SVI, effectively captures neighborhood-level risk for COVID-19–related hospitalizations, but its predictive power varies across different racial and ethnic groups, highlighting the possible need for measures that consider both socioeconomic deprivation and segregation.^38^ Research by Casillas et al. (2022), found that historical HOLC redlining measures were more closely associated with COVID-19 incidence and mortality rates at the census tract level in California than two other ABSMs which did not incorporate structural or historical racism.^13^ Importantly, variation in the time frame of studies can result in different conclusions about the association between ABSMs and the outcomes of interest. For the case of COVID-19, disparities were stronger in the earlier phases, and over time some disparities attenuated due to the high transmissibility of COVID-19 and other factors.

The association between ABSMs and other disease outcomes have been examined beyond COVID-19. For example, Bakhsh et al. assessed the relationship between tuberculosis (TB) incidence and ABSMs, including educational attainment, poverty thresholds, crowding, and the California HPI at the census tract level in California for 2012–2016.^39^ The authors found a clear inverse relationship between all the ABSMs and TB incidence, with areas in the lowest socioeconomic quartile for each ABSM experiencing TB incidence rates 2.0 to 3.6 times higher than areas in the highest socioeconomic quartile, across all ABSMs assessed.^39^ While we applaud the increasing use of systematic measures to address health disparities, the plethora of analytic options complicates the decision-making process for researchers and policymakers, who are left to choose ABSMs for use without clear guidance, often resulting in arbitrary selection.^16^

This study’s objective was to demonstrate a proof-of-concept approach to determine whether different ABSMs at the census tract level were more correlated with monthly public health outcomes, in this case COVID-19 incidence and mortality, in California. California was chosen due to its large and diverse population, significant variation in socioeconomic conditions across the state, and its status as an early epicenter of the COVID-19 pandemic, making it an ideal setting for examining disparities in health outcomes. We describe the design and structure of five different ABSMs: SVI, ADI, ICE, HOLC scores, as well as the California HPI which is a California-specific measure optimized to capture life expectancy at birth.^40–42^ Although many other measures exist, these five measures were chosen due to their relevance in California public health policy (HPI), common use in public health response (SVI, ADI), documented ability to account for racial/ethnic health disparities (ICE), and relevance to structural disadvantage rooted in historical disinvestment (HOLC grades). We constructed a census tract-level dataset of HOLC grades available in eight cities with HPI, ADI, SVI, and ICE measures for census tracts in those cities and also statewide. To ease comparison, we transformed SVI, ADI, and ICE to follow asset-based framing, which is used for HPI, where higher scores are interpreted as more opportunity or advantage. Census tracts were percentile ranked by their transformed scores and divided into quartiles, where the lowest (highest) quartile represents places with the least (most) opportunity for a healthy or advantaged life. We additionally assessed associations using Pearson’s correlation coefficient across the state of California. To assess temporal shifting in what ABSMs measure in urban landscapes, we further examined current ICE and HPI data with historical HOLC data for census tracts in Los Angeles (LA) city. By evaluating the most strongly associated ABSM with a health outcome month over month across a large geography, we aimed to highlight a process that could support public health planning, response, and evaluation, drawing insights on the extent such an approach may be suitable for a dynamically changing health-related landscape.

## Methods

### Data and Outcomes

We constructed two main datasets from various data sources. First, a census-tract-level dataset was constructed for California with publicly available data on HPI v3.0, ADI 2019, SVI 2020, ICE 2020, and HOLC grades – all based on census tracts delineated for the 2010 Census.^42–46^ HPI data was downloaded directly from the Public Health Alliance of Southern California’s HPI website; ADI, from the Neighborhood Atlas website administered by a research team at the University of Wisconsin’s Center for Health Disparities Research; SVI from the CDC/ATSDR website; and HOLC grades from the “Mapping Inequity: Redlining in New Deal America” website. HPI, SVI, ICE and HOLC grade datasets were available at the census tract level. Since ADI data was available at the census block level, an ADI census tract score was computed by averaging the ADI value for all blocks in a tract. The second dataset is a month-tract level dataset constructed from California’s COVID-19 surveillance data for the period February 2020 to December 2021, linked to the ABSM dataset. Counties were categorized by population size, with large counties defined as having populations greater than 106,000 and small counties having populations of 106,000 or fewer.

To make quantitative comparisons, ADI, SVI, and ICE measures for census tracts across California were transformed, percentile ranked, and then divided into quartiles and deciles, such that the directions of interpretation were similar to HPI. Thus, we refer to the 1^st^ quartile (or 1^st^ decile) as the 25% (or 10%) of California’s population who live in census tracts with the least opportunity for a healthy or advantaged life and the 4^th^ quartile (or 10^th^ decile), with the most opportunity.

Two main COVID-19 outcomes were constructed: monthly case rate and monthly mortality rate. Cases refer to individuals with a laboratory-confirmed, positive SARS-CoV-2 nucleic acid amplification test (NAAT; including polymerase chain reaction tests), as reported to the California Department of Public Health (CDPH). COVID-19 mortalities refer to individuals with confirmed COVID-19-associated deaths reported to the CDPH by county and city local health jurisdictions. Monthly rates are per 100,000 population at the census tract level over the study period. Census tracts had been assigned by CDPH to an individual’s residence as per addresses reported to the California Reportable Disease Information Exchange (CalREDIE). All population counts used in this study are 2019 ACS 5-year estimates.

### Analytic Approach

To conduct a comparative analysis of the five ABSMs, we first provide a qualitative overview of each of the five measures, including the year each measure was developed and the latest available data, their geographic unit, number of variables, and variable domains used to construct each ABSM. We briefly describe each ABSM’s design, notes for use in public health decision making, and a description of the transformation we applied to ADI, SVI, and ICE measures.

We then conducted descriptive statistics for tracts in California, large counties, small counties, and a city sample, which was constructed by excluding any tract that did not have a HOLC grade. By group, we presented demographic characteristics from 2019 ACS 5-year estimates; average HPI, transformed ADI, transformed SVI, and transformed ICE percentiles, including the frequency and proportion of tracts with missing ABSM data; proportion of tracts by each HOLC grade; and counties with tracts assigned HOLC grades. By group, we additionally present characteristics of each HPI constituent by HPI Policy Action area, given that the state of California and CDPH has recently utilized HPI for health equity focused policies.^47–49^ To quantitatively assess the ABSMs further, we employ various visualization and statistical techniques. First, we visualize maps of California and California regions with administrative boundaries to show the geographical distribution of tracts by ABSM deciles, including tracts that have missing ABSM data. For another perspective, we show how census tracts in each HPI quartile map to transformed ADI, SVI, and ICE quartiles (or missing ABSM data) by using an alluvial plot.

Correlation matrices are then constructed to quantify the strength of association across HPI and transformed ADI, SVI, and ICE percentiles, as well as quartiles statewide. Pearson correlation coefficients are used, which are defined as: 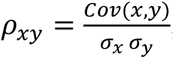, where *Cov*(*x*, *y*) is the covariance between ABSM percentile or quartile *x* and another ABSM percentile or quartile *y* among tracts in a sample, *σ*_*x*_ is the standard deviation of x, and *σ*_*y*_ is the standard deviation of y.

To identify the most correlated ABSM with monthly COVID-19 outcomes, we computed *ρ*_*xy*_ for the case rate or mortality rate of each month between February 2020 and December 2021 with the HPI, ADI, SVI, OR ICE percentile for tracts at a geography of interest (region, county, city). The maximum of the absolute values of all the correlation coefficients that were significant at the 10% level was selected as the most highly associated.

### Ethical Considerations

Analyses conducted were considered exempt from the State of California Health and Human Services Agency’s Committee for the Protection of Human Subjects (Project #2023-193), as data and results were considered essential components of California Department of Public Health public health surveillance.

### Data Statement

ABSM data utilized in in these analyses are available with analytic code at: https://github.com/Cesariddle/Index-Comparison-Analysis. ^50^ Monthly time-series data of COVID-19 outcomes at the census tract level are considered protected public health data. Investigators interested in accessing this data should contact the corresponding author to discuss the process for developing a data use agreement and accessing the data. All analyses were performed using Stata version 18 and R Statistical Software version 4.0.4 (R Group, Vienna Austria). For geospatial visualizations, we used the spmap package in Stata, and for alluvial plots, the tidyverse and ggalluvial packages in R were used.^51,52^

## Results

### Description of Area Based Socioeconomic Measures (ABSMs)

**Table 1** provides a qualitative description of the five selected ABSMs in raw form: California HPI version 3.0 (HPI v3.0), ADI 2019, SVI 2020, ICE 2020, and HOLC grades. Since new versions for the ABSMs are occasionally updated and refined, we note the version of the ABSM utilized in this study. The historical HOLC grades were the earliest to exist, with HPI and ICE initially developed more recently in 2014 and 2015, respectively. Data availability for the five ABSMs vary in terms of geographic scope and by level of geography unit. HPI v3.0 is California-specific (Utah is the only other state with a state-specific HPI as of time of writing) and, along with SVI and ICE measures, are made available at the county, census tract, and zip code tabulation area (ZCTA) levels. ADI is available at the census block level, and HOLC grades are only available at the tract and ZCTA levels for certain cities only. ADI, SVI, and ICE measures are available for all states in the US.

**Table 1.** Summary of area-based socioeconomic measures.

|  | California Healthy Places Index v3.0 (HPI) | Area Deprivation Index (ADI) 2019 | Social Vulnerability Index (SVI) 2020 | Index of Concentration at the Extremes (ICE) 2020 | Home Owners' Loan Corporation (HOLC) "Redlining" Grades |
| --- | --- | --- | --- | --- | --- |
| **Year initially developed** | 2014 | 2003 | 2011 | 2015 | 1935 |
| **Geographic Scope** | California only | All US states | All US states | All US states | Select cities only |
| **Geographic Unit** | County, Census Tract, Zip Code Tabulation Area | Census Blocks | County, Census Tract, Zip Code Tabulation Area | County, Census Tract, Zip Code Tabulation Area | Census Tract, Zip Code Tabulation Area |
| **Number of Variables** | 23 | 17 | 16 | 3 | 1 |
| **Variable Domains** | 1. Economic<br>2. Education<br>3. Social<br>4. Transportation<br>5. Health Care Access<br>6. Neighborhood<br>7. Housing<br>8. Clean Environment | 1. Education<br>2. Income/Employment<br>3. Housing<br>4. Household characteristics | 1. Socioeconomic Status<br>2. Household Characteristics<br>3. Racial and Ethnic Minority Status<br>4. Housing Type and Transportation | 1. Economic Inequality or Concentration of Wealth and Income by Race and Ethnicity Groups | 1. Desirability and risk for mortgage lending as graded by the US federal agency the Home Owners' Loan Corporation between the 1930-40s |
| **Race/Ethnicity Variables Included** | No | No | Yes | Yes | No |
| **ABSM Description & Interpretation** | Geographical units have HPI scores which can be percentile ranked, such that HPI percentile ranges from 0 to 100, with higher percentiles indicating areas with "the most opportunity to live a healthy life." Percentile rankings can also be divided into quartiles, where the lowest (highest) quartile refers to the "25% of communities with the least (most) opportunity to life a healthy life." | Geographic units are percentile ranked at the national level (0-100) and decile ranked at the state level (1-10). Lower (higher) values indicate areas with greater (lower) deprivation. | Geographic units are percentile ranked by overall SVI score as well as by themes. Both are sometimes subsequently divided further – for example, the top decile or "top 10%" are given a "flag" value" to indicate high vulnerability. Higher scores indicate areas with "greater vulnerability to environmental and public health hazards." | ICE is constructed by subtracting the number of people who belong to the "deprived extreme" (low-income black population) from the number of people belonging to the "privileged extreme" (high-income white population), divided by the total population of the geographic unit. ICE is a continuous variable that ranges from – 1 to 1, where – 1 corresponds with complete deprivation and 1 with complete privilege. | The HOLC used a four-grade system (A, B, C, D) to classify neighborhoods based on a range of factors, including housing stock, racial and ethnic composition, and perceived economic stability. Areas with grades A ("best") and B ("still desirable") were seen as "more desirable and less risky" for mortgage lending, while areas with grades C ("definitely declining") and D ("hazardous") were seen as "less desirable and riskier." |
| **ABSM Transformation for Comparative Analysis** | HPI serves as our reference: tracts are percentile ranked and split into quartiles, with lowest | Transform decile ranking into percentile and inverted the index so that the first 25 percentiles | Inverted the percentile ranking so that the first 25 percentiles indicate the | Transform the -1 to 1 index into percentile, the first 25 percentiles | No transformation applied. |

|  | quartile corresponding to the "least opportunity to live a healthy life" | indicate the most disadvantage / vulnerable | most disadvantage / vulnerable | indicate the most disadvantage / vulnerable |  |
| --- | --- | --- | --- | --- | --- |
| **Notes for Use** |  |  |  |  |  |
| *ABSM Maintenance & updates* | HPI is updated by the Public Health Alliance of Southern California with plans for national expansion. | ADI is updated by the "Neighborhood Atlas" at the University of Wisconsin's Center for Health Disparities Research. | SVI is updated by CDC/ATSDR and endorsed by the CDC at the national level and commonly referred to in national health recommendations. | Instructions on constructing ICE is available by the Public Health Disparities Geocoding Project 2.0 housed at Harvard University. | HOLC grades can be retrieved from the "Mapping Inequality: Redlining in New Deal America" website. |
| *ABSM construction & application notes* | HPI uses asset-based framing and maximizes the overall index's association with life expectancy at birth, so tied specifically to a health outcome. | Applying thresholds for ADI at the national versus state level may result in different conclusions. | More commonly utilized for public health emergency and disaster planning, preparedness and response related to natural disasters, environmental crises, and public health emergencies. | ICE can be used to identify areas of concentrated poverty and economic segregation, which can inform policy and advocacy efforts aimed at promoting economic equity and social justice. | Relevant for capturing historical discrimination and current health, socioeconomic, and educational inequalities. |
| **Sources** | *Public Health Alliance of Southern California, 2020* | *Kind and Buckingham, 2018; University of Wisconsin School of Medicine and Public Health, 2020.* | *Centers for Disease Control and Prevention Agency for Toxic Substances and Disease Registry, 2020.* | *Krieger, Chen, Waterman, 2020.* | *Fishback, LaVoice, 2021; Nelson et al. 2023 (version 3.0).* |

**Notes.**
HPI source: https://assets.website-files.com/613a633a3add5db901277f96/63320a9e98493bbdcc03d509_HPI3TechnicalReport2022-09-20.pdf
ADI source: https://www.neighborhoodatlas.medicine.wisc.edu/login
SVI source: https://www.atsdr.cdc.gov/placeandhealth/svi/img/pdf/Flanagan_2011_SVIforDisasterManagement-508.pdf
ICE source: https://www.hsph.harvard.edu/thegeocodingproject/covid-19-resources/
HOLC grades source: https://www.openicpsr.org/openicpsr/project/141121/version/V2/view

Developed by the Public Health Alliance of Southern California, HPI v3.0 is comprised of 23 variables, which fall into eight categories called Policy Action areas.^40,42,43,53^ A census tract’s HPI score is constructed from z-scores of each set of Policy Action area variables, which are then weighted to optimize prediction of life expectancy at birth. First described in an article published in 2003,^54^ADI was created by the US Health Resources and Services Administration and is updated by a research team at the University of Wisconsin. It contains 17 variables, constructed into a score with which census blocks are percentile ranked at the national level (0-100) and decile ranked at the state level (1-10) such that lower (higher) values indicate areas with greater (lower) deprivation.^55^ Given different ranking approaches, applying thresholds for ADI at the national versus state level may result in different conclusions. The CDC/ATSDR’s SVI has 16 variables, where geographic areas are percentile ranked by the overall score and by variable themes. To capture economic inequality or the concentration of wealth across race/ethnicity groups, ICE is developed from three variables by researchers at Harvard University. ICE’s interpretation is derived by subtracting the number of people who belong to the “deprived extreme” of wealth and income from the number of people belonging to the “privileged extreme”, divided by the total population of the geographic unit. In its original form, ICE is a continuous variable that ranges from -1 to 1, where -1 corresponds with complete deprivation and 1 with complete privilege. In this study, we report transformed values of ADI, SVI, and ICE such that the 1^st^ quartile (“q1”) refers to the 25% of places with the least opportunity for healthy or advantaged life, and the 4^th^ quartile (“q4”), the most opportunity. **Supplement A1** lists all the constituent variables for the five ABSMs.

Regarding cities, the HOLC used a four-grade system (A, B, C, D) to classify neighborhoods based on a range of factors, including housing stock, racial and ethnic composition, and perceived economic stability. Areas with grades A (“best”) and B (“still desirable”) were seen as “more desirable and less risky” for mortgage lending, while areas with grades C (“definitely declining”) and D (“hazardous”) were seen as “less desirable and riskier.“

In **Table 2**, we summarize characteristics of the census tracts in all of California’s 58 counties (using 2010 census tract boundaries), census tracts in large versus small counties (we define large as having a population greater than 106,000; otherwise, a county is small), and census tracts in our city analytic sample (i.e., any tract with a HOLC grade). Of California’s 8,057 census tracts, 97.3% (7,839) are in the 35 large counties with the remaining 2.7% (218) in the 23 small counties, and 13.9% (1,120) are in the city analytic sample. Across California, census tracts have an average estimated population of approximately 4,876 (standard deviation, SD: 2,281) with large county census tracts averaging a slightly higher population (4,893, SD: 2,283) and small county tracts averaging a slightly lower population (4,269, SD: 2,120). Using data from the ACS 2015-2019 5-year estimates, **Table 2** also provides averages for life expectancy at birth, and we observe varied composition of race/ethnicity groups distributed across counties by size, as well as with the city sample.^56^ The majority of census tracts in California have ABSM scores, with variation in missingness: 267 (3%) of the state’s census tracts are missing an HPI score, 139 (2%) are missing an ADI score, 1,248 (15%) are missing an SVI score, and 1,246 (15%) are missing an ICE score.

**Table 2.** Descriptive statistics of demographics and area-based socioeconomic measures for census tracts in California pooled, by county size, and city sample.

|  | A.<br>All Counties in California<br>(N=8,057 census tracts) |  |  | B.<br>Large County Sample<br>(N=7,839 census tracts) |  |  | C.<br>Small County Sample<br>(N=218 census tracts) |  |  | D.<br>City Sample<br>(N=1,120 census tracts) |  |  |
| --- | --- | --- | --- | --- | --- | --- | --- | --- | --- | --- | --- | --- |
|  | N | Mean or<br>Prop. | SD | N | Mean or<br>Prop. | SD | N | Mean or<br>Prop. | SD | N | Mean or<br>Prop. | SD |
| **Demographic Characteristics** |  |  |  |  |  |  |  |  |  |  |  |  |
| Population | 8,057 | 4,875.70 | 2,280.50 | 7,839 | 4,892.57 | 2,282.62 | 218 | 4,269.14 | 2,120.14 | 1,120 | 4,207.40 | 1,321.43 |
| Life expectancy at birth | 7,790 | 80.26 | 3.39 | 7,591 | 80.31 | 3.38 | 199 | 78.67 | 3.60 | 1,117 | 80.24 | 3.54 |
| Proportion of pop. White | 8,012 | 0.39 | 0.26 | 7,796 | 0.38 | 0.26 | 216 | 0.68 | 0.19 | 1,120 | 0.28 | 0.27 |
| Proportion of pop. Black | 8,012 | 0.06 | 0.09 | 7,796 | 0.06 | 0.09 | 216 | 0.02 | 0.03 | 1,120 | 0.09 | 0.13 |
| Proportion of pop. Asian | 8,012 | 0.14 | 0.16 | 7,796 | 0.14 | 0.16 | 216 | 0.03 | 0.05 | 1,120 | 0.15 | 0.16 |
| Proportion of pop. Native American | 8,012 | <0.01 | 0.02 | 7,796 | <0.01 | 0.02 | 216 | 0.02 | 0.04 | 1,120 | <0.01 | <0.01 |
| Proportion of pop. Pacific Islander | 8,012 | <0.01 | 0.01 | 7,796 | <0.01 | 0.01 | 216 | <0.01 | 0.01 | 1,120 | <0.01 | 0.01 |
| Proportion of pop. Multiple races | 8,012 | 0.03 | 0.02 | 7,796 | 0.03 | 0.02 | 216 | 0.03 | 0.02 | 1,120 | 0.03 | 0.02 |
| Proportion of pop. Other race | 8,012 | <0.01 | 0.01 | 7,796 | <0.01 | 0.01 | 216 | <0.01 | <0.01 | 1,120 | <0.01 | 0.01 |
| Proportion of pop. Latino/a | 8,012 | 0.38 | 0.26 | 7,796 | 0.39 | 0.27 | 216 | 0.22 | 0.17 | 1,120 | 0.44 | 0.30 |
| **Area-based socioeconomic measures** |  |  |  |  |  |  |  |  |  |  |  |  |
| Average HPI v3.0 percentile | 7,790 | 0.50 | 0.29 | 7,591 | 0.50 | 0.29 | 199 | 0.37 | 0.18 | 1,117 | 0.44 | 0.33 |
| Proportion Missing | 267 | 0.03 | - | 248 | 0.03 | - | 19 | 0.09 | - | 3 | <0.01 | - |
| Average ADI 2019 (transformed percentile) | 7,918 | 0.45 | 0.28 | 7,703 | 0.46 | 0.28 | 215 | 0.17 | 0.16 | 1,120 | 0.55 | 0.25 |
| Proportion Missing | 139 | 0.02 | - | 136 | 0.02 | - | 3 | 0.01 | - | 0 | 0.00 | - |
| Average SVI 2020 (transformed percentile) | 6,809 | 0.39 | 0.28 | 6,641 | 0.40 | 0.28 | 168 | 0.34 | 0.23 | 1,038 | 0.31 | 0.28 |
| Proportion Missing | 1,248 | 0.15 | - | 1,198 | 0.15 | - | 50 | 0.23 | - | 82 | 0.07 | - |
| Average ICE 2020 (transformed percentile) | 6,811 | 0.50 | 0.29 | 6,643 | 0.50 | 0.29 | 168 | 0.55 | 0.18 | 1,038 | 0.38 | 0.32 |
| Proportion Missing | 1,246 | 0.15 | - | 1,196 | 0.15 | - | 50 | 0.23 | - | 82 | 0.07 | - |
| Proportion by HOLC Grades |  |  |  |  |  |  |  |  |  |  |  |  |
| A (Best) | 48 | 0.01 | - | 48 | 0.01 | - | 0 | 0.00 | - | 48 | 0.04 | - |
| B (Still Desirable) | 280 | 0.03 | - | 280 | 0.03 | - | 0 | 0.00 | - | 280 | 0.25 | - |
| C (Definitely Declining) | 571 | 0.07 | - | 571 | 0.07 | - | 0 | 0.00 | - | 571 | 0.51 | - |
| D (Hazardous) | 221 | 0.03 | - | 221 | 0.03 | - | 0 | 0.00 | - | 221 | 0.20 | - |
| Proportion Missing | 6,937 | 0.86 | - | 6,937 | 0.86 | - | 0 | 0.00 | - | 0 | 0.00 | - |
| County of Census Tract with HOLC Grades |  |  |  |  |  |  |  |  |  |  |  |  |
| Alameda | 120 | 0.11 | - | 120 | 0.11 | - | 0 | 0.00 | - | 120 | 0.11 | - |
| Fresno | 9 | 0.01 | - | 9 | 0.01 | - | 0 | 0.00 | - | 9 | 0.01 | - |
| Los Angeles | 794 | 0.71 | - | 794 | 0.71 | - | 0 | 0.00 | - | 794 | 0.71 | - |
| Sacramento | 9 | 0.01 | - | 9 | 0.01 | - | 0 | 0.00 | - | 9 | 0.01 | - |
| San Diego | 76 | 0.07 | - | 76 | 0.07 | - | 0 | 0.00 | - | 76 | 0.07 | - |
| San Francisco | 95 | 0.08 | - | 95 | 0.08 | - | 0 | 0.00 | - | 95 | 0.08 | - |
| San Joaquin | 3 | <0.01 | - | 3 | <0.01 | - | 0 | 0.00 | - | 3 | <0.01 | - |
| Santa Clara | 14 | 0.01 | - | 14 | 0.01 | - | 0 | 0.00 | - | 14 | 0.01 | - |

**Notes:** Large counties are California counties with population >106,000; small counties have populations with 106,000 or fewer. The city sample includes any tract with a HOLC score. Prop is proportion; SD is standard deviation. Population; life expectancy at birth; proportions by race and ethnicity are from ACS 2015-2019 5-year estimates. HPI is California Healthy Places Index, version 3.0. ADI is Area Deprivation Index. SVI is Social Vulnerability Index. ICE is Index of Concentration at the Extremes. HOLC is Home Owners' Loan Corporation. Census tracts are based on the boundaries delineated in the 2010 US Census.

Within the city analytic sample, the 1,120 census tracts with HOLC grades are entirely in large counties. Among census tracts in the city analytic sample, 4% have Grade A; 25%, Grade B; 51%, Grade C; and 20%, Grade D. Further, 71% of these census tracts are in Los Angeles County; 11%, in Alameda County; 8%, in San Francisco County; and 7%, in San Diego County (**panel D).**

We created state and regional choropleth maps to show how California census tracts grouped by deciles according to ABSMs with statewide data are geographically distributed (see **Supplements A2 and A3**). There are general patterns noted by areas with high population density (smaller census tracts), areas with low population density (large census tracts), and areas with missing ABSM data.

In the first year of the COVID-19 pandemic, the state of California and the California Department of Public Health used HPI v2.0 to anchor its pandemic reopening and response strategy in health equity.^47,48^ Because of the statewide use of HPI, we present descriptive statistics of each of the 23 HPI constituents in **Supplement A4** for all of California’s 58 counties, tracts in large versus small counties, and tracts in our city sample. We note that census tracts in small counties are on average worse off in terms of all HPI constituents in economic and education Policy Action areas but have the same average rate of healthcare access (89% for large and small counties; 85% for the city sample).

### Area-Based Socioeconomic Measures across California

**Figure 1** depicts an alluvial plot of how California’s 8,057 census tracts grouped by their HPI quartile map to quartiles of transformed ADI, SVI, and ICE measures. Based on thickness of bands flowing from one measure to another, we observe that census tracts falling into the least and most advantaged quartiles (q1 and q4, respectively) appear more likely to maintain that quartile across ABSMs (thicker bands) than tracts falling into q2 and q3 across the ABSMs (thinner bands). The tabulated frequencies of tracts across these quartiles confirm this (see **Supplement A5**). SVI and ICE are most similar in terms of tracts falling into q1 and q4 for both ABSMs. ADI and ICE appear to be the least similar in that sense. Since HOLC grades are available for a smaller subset of the state’s census tracts, they are not incorporated into the alluvial plot.

**Figure 1.**
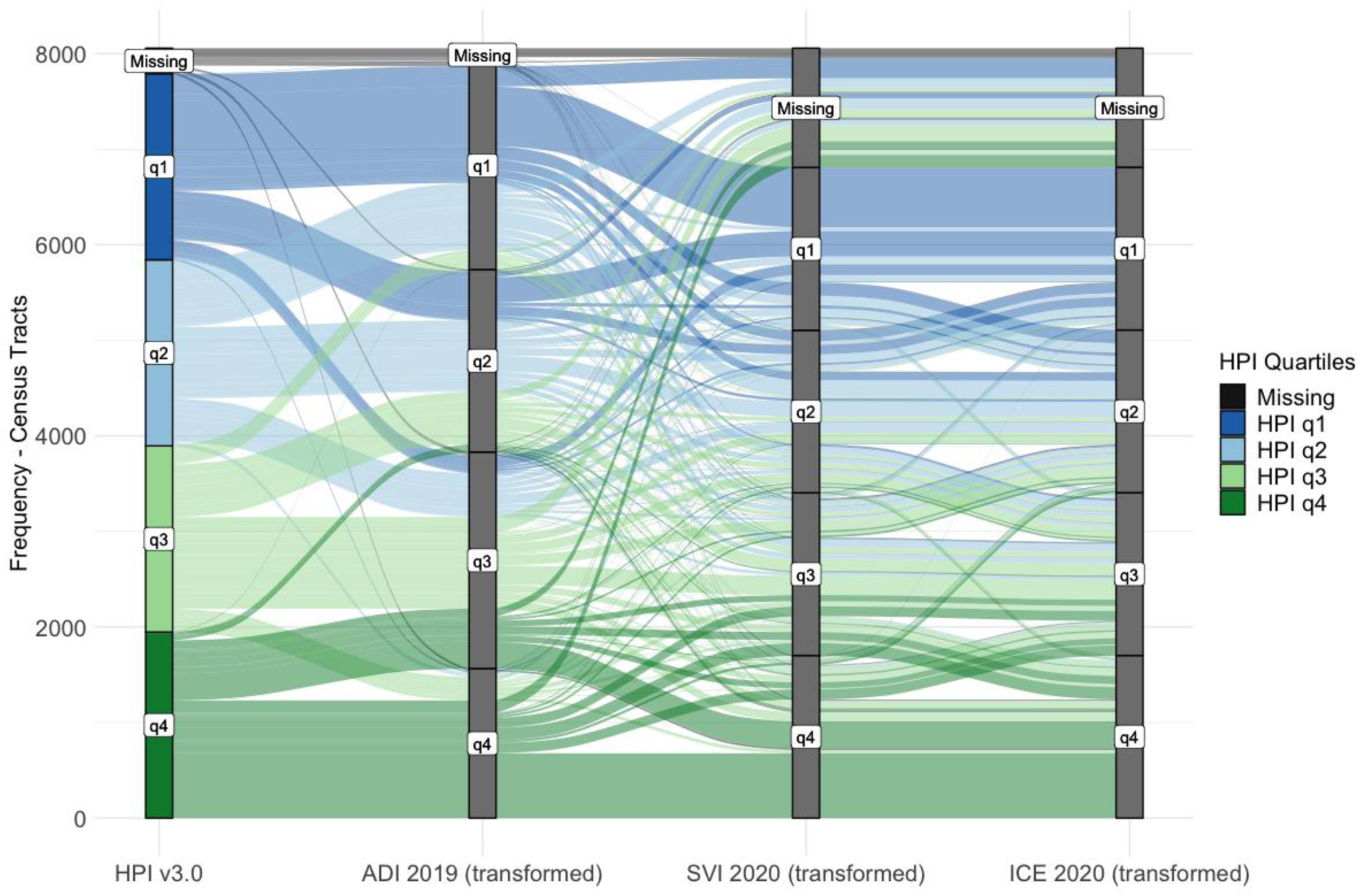
Alluvial plot demonstrating relationship of Californiás census tracts by quartiles mapped to HPI and transformed ADI, SVI and ICE. Notes: HPI is California Healthy Places Index, version 3.0. ADI is Area Deprivation Index. SVI is Social Vulnerability Index. ICE is Index of Concentration at the Extremes. ADI, SVI, and ICE are transformed as described in Table 1. Labels “q1” to “q4” refer to quartile 1 to quartile 4 for corresponding measure, with HPI q1 (2,3,4) referring to HPI quartile 1 (2,3,4). This plot demonstrates that by large, census tracts at the most and least advantage groups are similarity categorized by these indices whereas there are more dynamic shifts between categories of advantage in the middle. Census tracts are based on the boundaries delineated in the 2010 US Census. Census tracts with missing data were included to see the extent of missingness across different ABSMs.

Correlation matrices of the four statewide ABSMs demonstrate significant correlations with each other (**Figure 2a**). SVI and ICE, SVI and HPI, and ICE and HPI have correlation coefficients greater than 0.6 indicative of very strong positive correlations. A positive (negative) value means the two ABSMs are positively (negatively) correlated. Overall, these correlations suggest that the ABSMs may measure similar SDOH or contextual dynamics, such as socioeconomic status, racial segregation, and environmental conditions, that contribute to the variation in these ABSMs. The correlation matrix across quartiles in **Figure 2b** show how correlations between the indices at the first and fourth quartiles are significant and the strongest, with all the correlation coefficients greater than 0.6 falling into this category, suggesting that these ABSMs are more correlated in how they identify communities of most and least advantage or opportunity. For example, quartile pairs with correlation coefficients greater than 0.6 include: HPIq1 with q1s of SVI and ICE; HPIq4 with q4s of SVI, ICE, and ADI; and SVIq1 with ICEq1. Conversely, some pairs of second and third quartiles show lower, but significant correlation coefficients (e.g., HPI’s q2 and q3 with ADI q2 and q3), suggesting that communities not in the most and least advantaged groups may be categorized slightly differently based on how each ABSM captures intermediate levels of socioeconomic disadvantage or vulnerability. A few pairs are not correlated at all (e.g., ADI q3 with SVIq2 and ICEq2) at the 10% significance level, suggesting that these measures may capture different aspects of SDOH in these middle categories.

**Figure 2.**
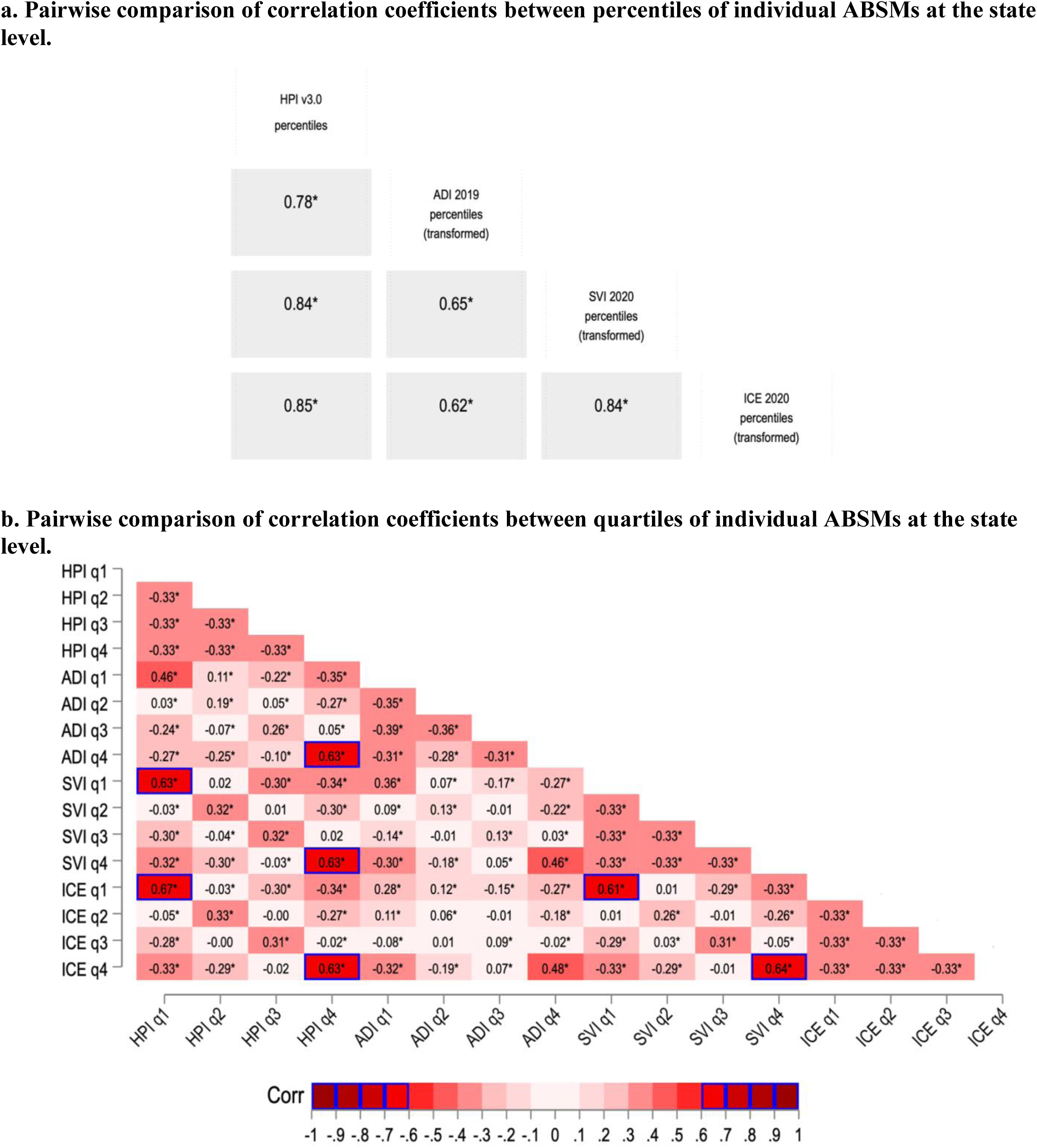
Correlation matrices among statewide ABSMs percentiles and quartiles. **Notes:** Values in each box are pairwise Pearson correlations (Corr) with significance designated as: * *p*<0.1. HPI is California Healthy Places Index, version 3.0. ADI is Area Deprivation Index. SVI is Social Vulnerability Index. ICE is Index of Concentration at the Extremes. ADI, SVI, and ICE are transformed as described in Table 1. Labels “q1” to “q4” refer to quartile 1 to quartile 4 for corresponding measure, with HPI q1 (2,3,4) referring to HPI quartile 1 (2,3,4). Blue boxes represent a correlation value greater than 0.60. In this context, a positive value reflects a positive correlation, meaning that as one value rises, so does the other. A negative value reflects a negative correlation, meaning as one value increases, the other decreases.

### ABSMs across cities

We next explored relationships between ABSMs and HOLC grades with our city analytic sample. We first examined city census tracts in Los Angeles County, the most populated county in the US. Los Angeles county accounts for 29% of California’s census tracts and almost 26% of the state population. We focus particularly on Los Angeles city because it represents 71% of census tracts with HOLC data. **Figure 3a** is a map of the locations of Los Angeles city census tracts assigned a HOLC grade (N=794, 34%). A scatterplot of these 794 census tracts based on their HPI and ICE percentiles with colors designating HOLC grades is depicted in **Figure 3b**. The 45-degree line marks the values at which the HPI percentile equals the ICE percentile. The dotted lines corresponding to HPI and ICE delineate where places with the least (red) and most (green) opportunity for a healthy or advantaged life fall. We observed a strong positive correlation between HPI and ICE percentiles. The table in **Figure 2b** shows the percentage of census tracts in each HOLC grade (A, B, C, D) that fall within the first quartiles of both HPI and ICE and the percentage falling in the 4th quartile of both indices. Notably, 93 (59.6%) of the 156 tracts previously assigned Grade D fall in the 1st quartiles for both HPI and ICE, whereas a smaller concentration of Grade D census tracts (only 6, 3.8%) fall in the 4^th^ quartiles for both HPI and ICE. On the other end of the spectrum, 20 (62.5%) of the 32 census tracts assigned Grade A are in both ABSMs’ 4th quartiles, and no Grade A census tracts fall in both 1st quartiles. **Supplement A6** includes choropleth maps of Los Angeles County by HPI and transformed ADI, SVI, and ICE percentiles grouped by deciles.

**Figure 3.**
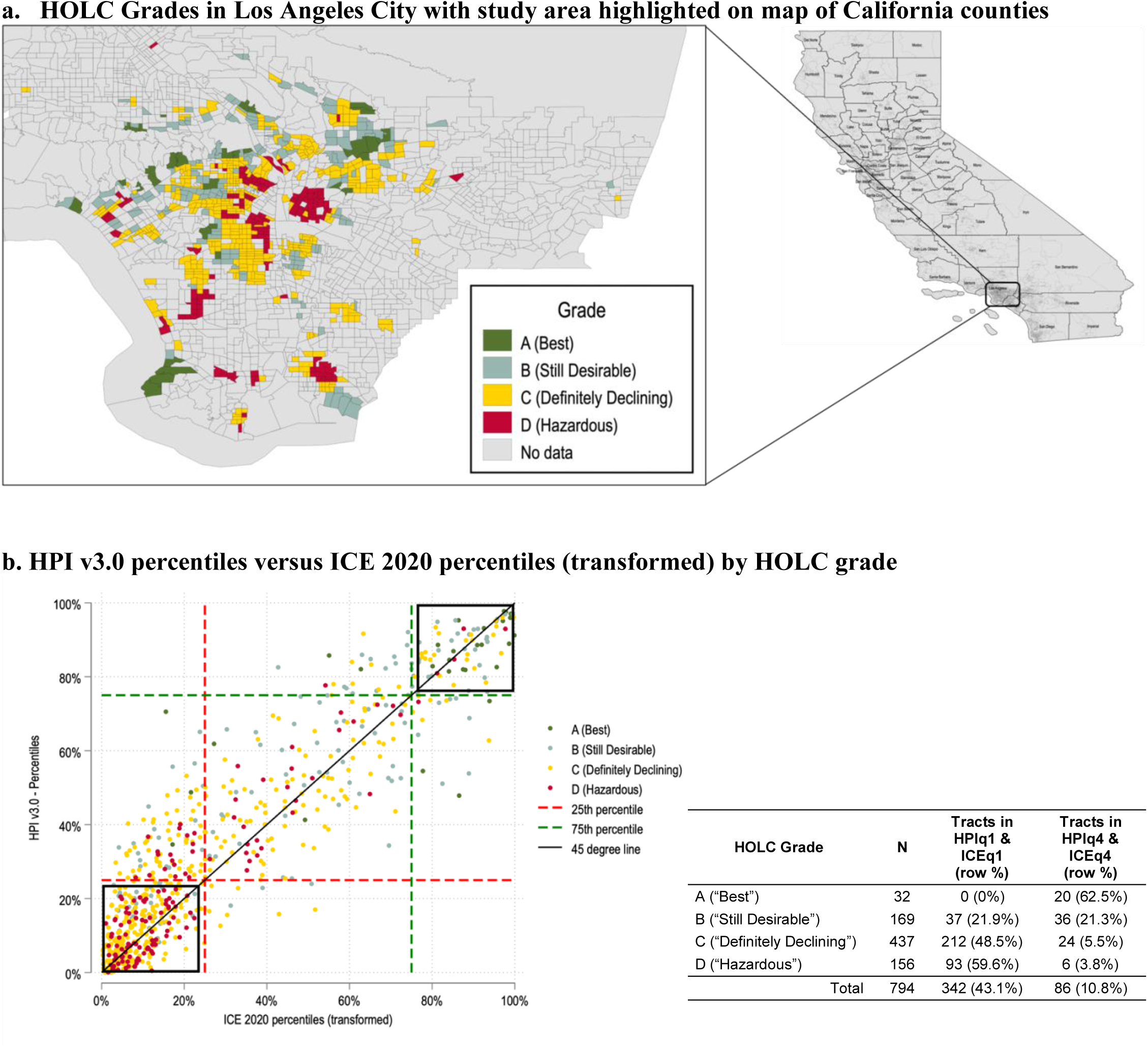
Comparison of present-day ABSM with historical HOLC grades for Los Angeles City census tracts (N=794) **Notes:** HOLC grades are Home Owners’ Loan Corporation’s assigned grades from the 1930s for areas corresponding to census tracts. Census tracts are based on the boundaries delineated in the 2010 US Census. Areas with grades A (“best”) and B (“still desirable”) were seen as “more desirable and less risky” for mortgage lending, while areas with grades C (“definitely declining”) and D (“hazardous”) were seen as “less desirable and riskier.” HPI is California Healthy Places Index, version 3.0. ICE is Index of Concentration at the Extremes, transformed as described in Table 1. The map in (a) displays the 794 census tracts in Los Angeles City with colors denoting tracts with available HOLC grades and ABSM data. The scatterplot in (b) shows HPI versus ICE percentiles, with points color coded by HOLC grades, a black box on the lower left highlighting areas in the lowest 25th percentile (most disadvantaged) for both ABSMs, a black box on the upper right highlighting areas in the highest 75th percentile (most advantaged), red dashed lines representing the HPI and ICE 25th percentile, green dashed lines representing HPI and ICE 25^th^ percentiles, and 45-degree black line denotes the values at which HPI percentile equals ICE percentile. Census tracts identified as least advantaged by contemporary ABSMs are by large the same communities that reflect historic disinvestment as demonstrated with HOLC grades as a proxy. We focus on Los Angeles city, because it represents 71% of census tracts with HOLC data.

### Relationships between ABSMs and monthly COVID-19 outcomes by region, county, and city

We explored the correlations between ABSM scores and monthly COVID-19 case and mortality rates at multiple geographic levels. Statewide results are presented in **Figure 4** by region and county (4a and 4b) and for the city sample (4c and 4d). Regions refer to the five California public health regions as determined by the CDPH and the California Health and Human Services Agency.^57^ Across the state, nearly all census tracts for each region and county were included in the analysis. For each timepoint, the value in each cell represents the correlation coefficient between the most strongly associated ABSM and the monthly COVID-19 outcome, as long as the correlation is significant at the 10% level. HPI is the ABSM most associated with monthly case rates for the Bay Area and Southern California regions (**Figure 4a**). Although case rates across months for different counties within the Bay Area region were most associated with HPI, HPI’s relationship to the Southern California region’s monthly case rates appear to be driven by Los Angeles and Orange counties. Monthly case rates in other counties in Southern California, such as Riverside, San Bernardino, San Luis Obispo, and Ventura counties, appear to be most associated with ICE. Three other general patterns exist. First, in 2020 and 2021, several months showed no significant association between case rates and ABSMs at the 10% significance level. This trend is most prominent in counties in the Northern California and Greater Sacramento regions. Some counties lacked COVID-19 data, interpreted as no reported cases, some extremely low-population counties like Alpine and Sierra counties, had no calculated correlations due to the presence of only a single census tract – thus lacking any variation in ABSM and COVID-19 outcome data. Finally, across some regions and counties, monthly case rates appear to become most associated with ADI after March 2021, corresponding to when vaccines became available to the general population. ADI was rarely noted as the most associated ABSM prior to that time point, except for a small number of counties, such as Yolo County.

**Figure 4.**
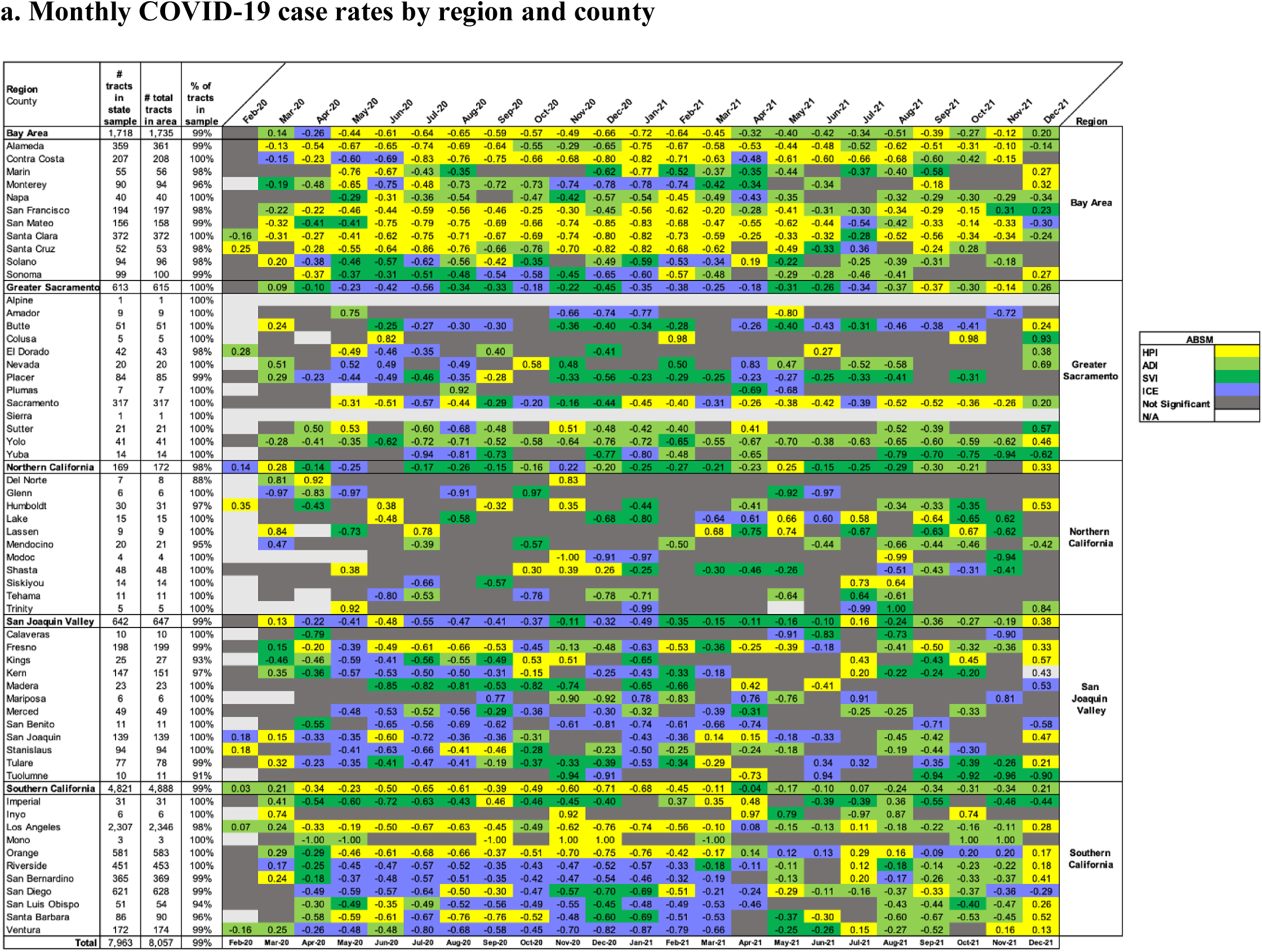
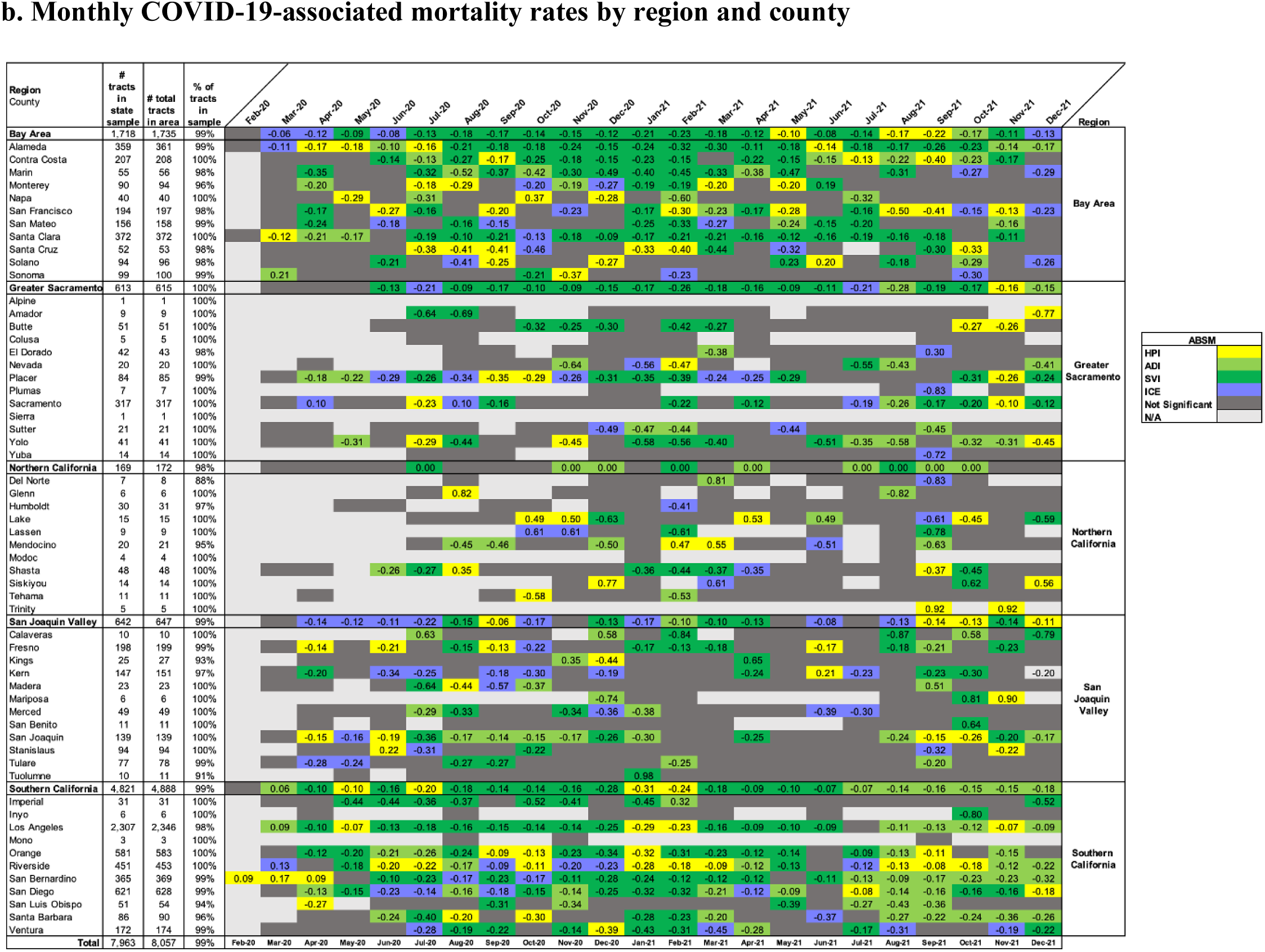
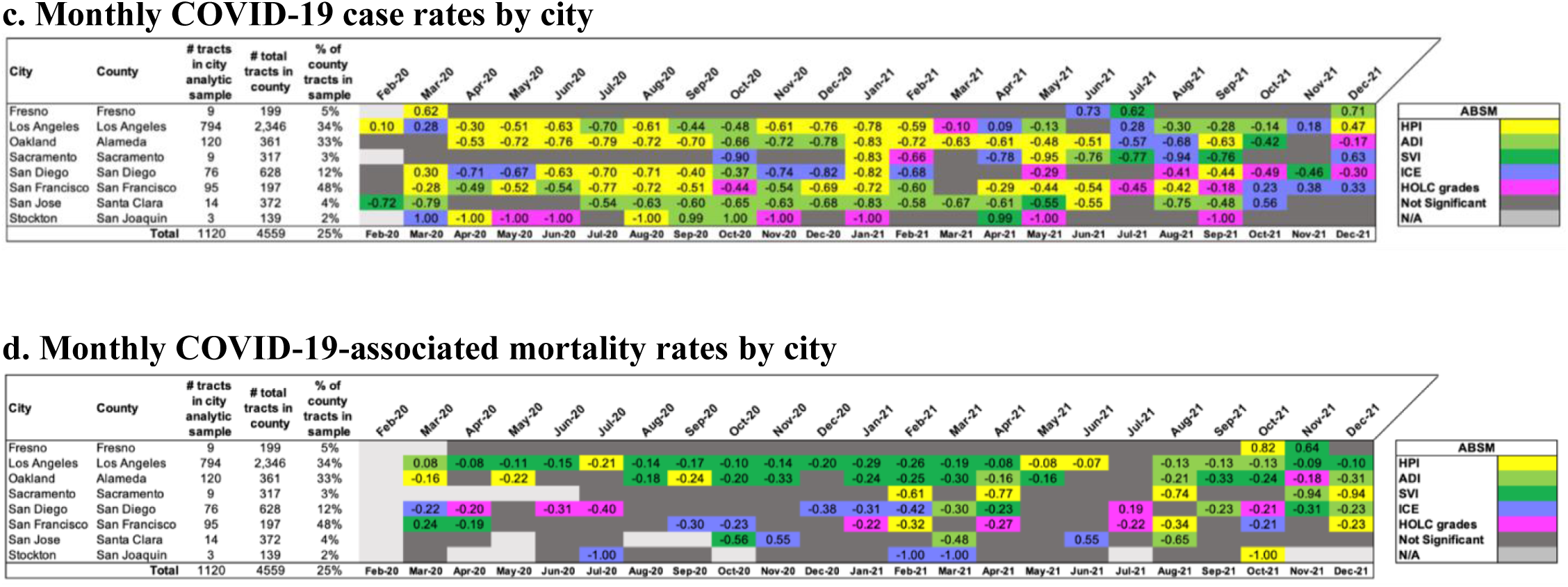
Most correlated ABSM with monthly COVID-19 outcomes by region, county, and city: February 2020 to December 2021. **Notes:** The value in each box is the pairwise Pearson correlation of the most correlated ABSM with the COVID-19 outcome for that month and geographical unit, as long as the *p-*value<0.1. When a box does not have a value, either no ABSM was correlated at the 10% significance level (dark grey designating “Not Significant”) or there was no variation in the COVID-19 outcome (light grey designating “N/A” as not applicable). HPI is California Healthy Places Index, version 3.0. ADI is Area Deprivation Index. SVI is Social Vulnerability Index. ICE is Index of Concentration at the Extremes. ADI, SVI, and ICE are transformed as described in Table 1. HOLC grades refers to the Home Owners’ Loan Corporation’s grades. Negative values indicate a negative correlation between ABSM scores and COVID-19 outcomes of interest (Case and Mortality Rate). Positive values indicate a positive correlation between ABSM scores and COVID-19 outcomes. For comparison, HOLC grade A was recategorized as 1, grade B as 2, grade C as 3 and grade D as 4. Census tracts are based on the boundaries delineated in the 2010 US Census.

In monthly COVID-19 mortality rate data shown in **Figure 4b**, data limitations impacted the ability to draw meaningful and significant conclusions regarding any relationship between mortality with ABSMs during 2020. However, for the Bay Area and Southern California regions, which are more contiguously populated than the other regions and have more tracts, we observe various patterns. For the Bay Area, SVI is most prominently associated with mortality across the region, month over month, with the exception of the beginning of the pandemic in 2020 and after August 2021. This appears to be driven by Alameda, Contra Costa, and Santa Clara counties, which are the three counties with the greatest number of census tracts. This observation from both Tables 2a and 2b suggests that regional associations are often influenced by large population counties, indicating that the use of ABSMs might be more effective at the state or county level rather than the regional level.

Within the city sample which includes HOLC grades, HPI appears to be the most prominent ABSM associated with monthly case rates, particularly for LA, Oakland, San Diego, and San Francisco cities (**Figure 4c**). In contrast, San Jose’s monthly case rates most frequently have the strongest association with ADI. We found this to be particularly the case until the summer months of 2021. Notably, a significant ABSM associated with case rates for Fresno, Sacramento, and Stockton, which are the three cities with the fewest number of census tracts (9, 9, and 3, respectively), was difficult to identify with our process. When examining mortality rates, many months across the cities had outcomes that were not significantly associated with the monthly outcome (**Figure 4d**). This was not the case for Los Angeles city, whose mortality rates were most associated with ADI, particularly until April 2021.

## Discussion

By comparing various ABSMs, we offer evidence-based insights on integrating ABSMs for public health use, as well as a proof-of-concept approach for identifying and addressing public health disparities. First, our analysis demonstrates, not surprisingly, that these ABSMs are quite correlated with one another, particularly for ABSMs that have similar constituent determinants and which utilize similar data sources. These similarities most often manifest among communities falling in quartiles experiencing the most or least advantage/opportunity. Where these ABSMs diverge is in describing communities in the middle quartiles of advantage/opportunity, suggesting that the differences in underlying data or constituent determinants for ABSMs creates more nuance in identifying communities not in the most and least advantaged groups.

Common to all the ABSMs we included in our study was their limited utility in contexts for which data is limited, such as community-level data on social or structural determinants in a low population setting, or public health surveillance data for rare health outcomes. This has at least two large implications for future ABSM use in public health policy and program planning. First, using ABSMs to inform public health decision-making or evaluation requires not only meaningful data but data in an adequate volume with meaningful resolution, meaning either at the individual level, at sufficiently specific geographic levels, and with meaningful temporal resolution. Those interested in applying ABSMs should consider the availability and type of data necessary to implement these tools in their area of interest. To explore this further, we generate correlation matrices of statewide ABSM quartiles, similar to Figure 2b, but for census tracts within large counties (i.e., with populations greater than 106,000) or small counties separately (**Supplement A7**). The associations within ABSM quartiles remained robust among census tracts in the 35 large counties, which account for 97.3% of the state’s 8,057 census tracts and 97.6% of the state’s population (**A6a**). In contrast, associations were not as significant among tracts in the 23 small population counties, which account for 2.7% of the state’s tracts and 2.4% of the state’s population (**A6b**). At the census tract-level, we also observed that these small-population areas were likely to lack the necessary amount of social, demographic, economic, environmental, health, and other data required to construct an ABSM from constituent data sources. This small-area estimation challenge is not unique to ABSMs and is consistent with challenges in constructing COVID-19 hospitalization forecasting models, as reported in a recent study on California.^57^

Since insufficient sample sizes may inhibit the derivation of substantive quantitative insights that can be drawn with samples boasting larger sample sizes, future efforts using ABSMs to address health disparities should carefully consider additional or alternative approaches to identify need in small population areas to prevent inequitable exclusion of these populations. While some studies have described approaches taken to develop small-area indices, such as in France, Denmark, and Spain,^58^ those approaches are not yet commonly applied to the US. A more recent implementation did occur in California with the development of the California Vaccine Equity Metric (VEM), which was based on the California HPI and for distributing COVID-19 vaccines during the pandemic. The VEM involved imputing data for ZCTAs lacking sufficient measures to construct an HPI score, demonstrating a practical approach to address data limitations in public health planning.^59^ Further, qualitative research methods and community engagement and participation approaches for research can provide valuable insights into local needs and contextualized resource allocation.

Second, although ABSM percentiles were highly correlated, they were not perfectly correlated. What may matter more is not when ABSMs are similar but when they result in different conclusions about the health and well-being of a community. We agreed with previous studies that measures for directing resources and prioritizing populations should be customized for specific public health goals and objectives.^14^ Our study presents one retrospective approach as a proof-of-concept for identifying the most fit-for-purpose ABSM for public health outcomes across geographical areas and over time, in this case using correlation matrices to describe associations between COVID-19 outcomes across California during the height of the pandemic. Future research should examine whether other approaches can be more useful in real-time or near-real time. Future exploration may also want to consider validating thresholds for measures associated with disease outcomes, integrating community-identified factors, as well as multiple hypotheses testing to quantitatively and qualitatively understand how ABSMs as place-based factors and diseases can improve targeting public health efforts.

The selection of ABSMs has important implications not only for identifying communities in need but also for accurately capturing health disparities when utilized at a high level of data resolution. For example, Moss et al. (2021), in a large-scale national analysis, assessed the validity of census tract and county-level ABSM indicators as proxies for individual-level socioeconomic status (SES), such as household income, poverty status, and educational attainment. The authors found moderate but imperfect concordance between individual-level SES indicators and their respective place-based measures. Notably, census tract-level ABSMs demonstrated greater precision compared to corresponding county-level measures when approximating individual SES.^60^ Their findings suggest potential risks of misclassification and attenuation in area-based analyses highlighting the importance of ABSM selection and caution when interpreting associations between ABSMs and health outcomes.

However, some insights are relevant for planning from our retrospective analysis. When we considered the relationship between these ABSMs and COVID-19 outcomes across different levels of geographies, we observed varying associations. For case rates, HPI stood out as the most prominent ABSM associated with outcomes month over month in the Bay Area. Our findings in the Bay Area are consistent with another study that reported higher social vulnerability, as defined by the CDC’s SVI, was associated with higher chances of testing positive for COVID-19.^61^ In the Greater Sacramento region, ICE demonstrated a better correlational measure with COVID-19 outcomes, suggesting that this measure may be more effective at capturing the underlying socioeconomic disparities that influence COVID-19 outcomes in this region. This implies that policies aimed at reducing these disparities might benefit from focusing on the factors ICE measures, such as income inequality and segregation, to better address vulnerabilities that exacerbate the impact of pandemics. Northern California exhibited a stronger association with SVI, while Southern California, including the San Joaquin Valley, showed higher correlations with ICE. These findings, consistent with findings from other studies on social vulnerability and racial health disparities for COVID-19 testing, hospitalization, and mortality, suggest that geographical differences across and within the regions may be influencing these patterns.^61^

Notably, when possible, the epidemiology of a disease or the drivers of a health issue of interest should be considered when selecting an ABSM, as it can aid in defining and addressing populations in need. It may be crucial to assess which ABSMs appropriately capture drivers of outcomes of interest. For example, an ABSM lacking environmental indicators might not be suitable for asthma where environmental factors drive outcomes. Future research is warranted on developing robust evidence on how effective ABSMs are for identifying priority populations and targeting resources across diverse health areas.

It is worth noting that the historical and ongoing interplay among socioeconomic status, race/ethnicity, social environment, and health in the US makes it difficult to distinguish what contributes to health disparities. Addressing disparities from disproportionate disease burdens across racial/ethnic groups is an ongoing challenge. Regardless of data limitations or availability, the use of race/ethnicity variables as an underlying framework for policies or program design in the state of California is not permitted, and similar restrictions may apply elsewhere.^62^ This makes the use of ABSMs such as SVI and ICE, which embed race/ethnicity variables, and measures related to historical disinvestment and structural racism, like HOLC grades, potentially challenging to implement as the basis for policy. Despite this, we have found that the correlation between ABSMs that include and exclude race/ethnicity suggest these measures capture similar social and economic factors. This implies that policies can still effectively target areas of need using ABSMs without explicitly including race/ethnicity variables. A further limitation is the substantial time investment required for detailed assessments of ABSMs. Public health professionals may not have the capacity to perform these analyses when an ABSM is urgently needed, especially in rapidly emerging health threats. Furthermore, when planning for new or evolving health priorities, such as novel infectious disease variants, retrospective data may not be available. In these cases, it may be practical to leverage data from analogous, well-documented conditions to guide ABSM application, enabling practitioners to make informed choices in the absence of condition-specific data.

Our results suggest variation in the associations between ABSMs and COVID-19 outcomes over time. Evidence shows that California’s use of HPI in equitably allocating vaccines statewide in March 2021 reduced disparities in these outcomes, but did not eliminate them.^49,59^ This suggests that during the pandemic, a more geographically and time-sensitive approach may have been more effective than a one-size-fits-all strategy for identifying populations to address health disparities. Future research may explore whether an alternative approach could have been more effective. Additionally, considering meaningful temporal resolution is critical, as disparities can change over time because of factors like transmissibility, natural immunity, vaccinations, and changing public health measures. This underscores the importance of using models that account for spatial and temporal variation. Whether this approach would be effective for other health conditions requires careful consideration of geographical areas with limited or missing data, and other contextual factors not easily measured.

## Data Availability

ABSM data utilized in in these analyses are available with analytic code at: https://github.com/Cesariddle/Index-Comparison-Analysis. Monthly time-series data of COVID-19 outcomes at the census tract level are considered protected public health data. Investigators interested in accessing this data should contact the corresponding author to discuss the process for developing a data use agreement and accessing the data.

## Acknowledgments

The authors would like to thank the California Department of Public Health staff who participated in seminars and meetings for their valuable support and feedback. We would also like to specifically acknowledge Tomás León, Lauren White, and Jason Vargo for their helpful contributions throughout the development of this manuscript.

## Author Contributions

CAG – Data curation, Formal analysis, Investigation, Methodology, Visualization, Writing – review and editing.

ATK – Conceptualization, Data curation, Formal analysis, Investigation, Methodology, Supervision, Validation, Visualization, Writing – original draft, Writing – review and editing.

PBS – Conceptualization, Funding acquisition, Investigation, Methodology, Supervision, Validation, Writing – review and editing.

## References

1. Marmot, M. & Allen, J. Prioritizing health equity. Health in All Policies 63, (2013).

2. Horwitz, L. I., Chang, C., Arcilla, H. N. & Knickman, J. R. Quantifying Health Systems’ Investment In Social Determinants Of Health, By Sector, 2017–19: Study analyzes the extent to which US health systems are directly investing in community programs to address social determinants of health. Health Affairs 39, 192–198 (2020).

3. Mays, V. M., Ponce, N. A., Washington, D. L. & Cochran, S. D. Classification of race and ethnicity: implications for public health. Annual review of public health 24, 83–110 (2003).

4. Dressler, W. W., Oths, K. S. & Gravlee, C. C. Race and ethnicity in public health research: models to explain health disparities. Annu. Rev. Anthropol. 34, 231–252 (2005).

5. Kawachi, I., Daniels, N. & Robinson, D. E. Health disparities by race and class: why both matter. Health Affairs 24, 343–352 (2005).

6. Braveman, P. A., Cubbin, C., Egerter, S., Williams, D. R. & Pamuk, E. Socioeconomic disparities in health in the United States: what the patterns tell us. American journal of public health 100, S186–S196 (2010).

7. Barr, D. A. Health Disparities in the United States: Social Class, Race, Ethnicity, and Health. (JHU Press, 2014).

8. World Health Organization & others. Operational framework for monitoring social determinants of health equity. (2024).

9. Rehkopf, D. H. et al. Monitoring socioeconomic disparities in death: comparing individual-level education and area-based socioeconomic measures. American journal of public health 96, 2135–2138 (2006).

10. Krieger, N. et al. Choosing area based socioeconomic measures to monitor social inequalities in low birth weight and childhood lead poisoning: The Public Health Disparities Geocoding Project (US). Journal of Epidemiology & Community Health 57, 186–199 (2003).

11. Phillips, R. L. et al. How other countries use deprivation indices—and why the United States desperately needs one. Health Affairs 35, 1991–1998 (2016).

12. Trinidad, S. et al. Use Of Area-Based Socioeconomic Deprivation Indices: A Scoping Review And Qualitative Analysis: Study examines socioeconomic deprivation indices. Health Affairs 41, 1804–1811 (2022).

13. Casillas, E. et al. The Color of a Pandemic: The Association Between Historical Residential Redlining and COVID-19 Outcomes in California. in C14. Burnout, Disparities, and Outcomes of the COVID-19 Pandemic A3693–A3693 (American Thoracic Society, 2022).

14. Ong, J. D. & Ong, P. M. Vulnerability Indicators and At-Risk Smaller Populations in California and Los Angeles. (2021).

15. Kaalund, K., Thoumi, A., Bhavsar, N. A., Labrador, A. & Cholera, R. Assessment of Population-Level Disadvantage Indices to Inform Equitable Health Policy. The Milbank Quarterly (2022).

16. Tipirneni, R., Schmidt, H., Lantz, P. M. & Karmakar, M. Associations of 4 Geographic Social Vulnerability Indices With US COVID-19 Incidence and Mortality. American Journal of Public Health 112, 1584–1588 (2022).

17. Deziel, N. C. et al. Assessing community-level exposure to social vulnerability and isolation: Spatial patterning and urban-rural differences. Journal of exposure science & environmental epidemiology 33, 198–206 (2023).

18. Tate, E. Social vulnerability indices: a comparative assessment using uncertainty and sensitivity analysis. Natural Hazards 63, 325–347 (2012).

19. Kind, A. J. & Buckingham, W. R. Making neighborhood-disadvantage metrics accessible— the neighborhood atlas. The New England journal of medicine 378, 2456 (2018).

20. Flanagan, B. E., Gregory, E. W., Hallisey, E. J., Heitgerd, J. L. & Lewis, B. A social vulnerability index for disaster management. Journal of homeland security and emergency management 8, (2011).

21. Krieger, N., Chen, J. T., Waterman, P. D., Rehkopf, D. H. & Subramanian, S. Race/ethnicity, gender, and monitoring socioeconomic gradients in health: a comparison of area-based socioeconomic measures—the public health disparities geocoding project. American journal of public health 93, 1655–1671 (2003).

22. Fishback, P. V., LaVoice, J., Shertzer, A. & Walsh, R. The HOLC Maps: How Race and Poverty Influenced Real Estate Professionals’ Evaluation of Lending Risk in the 1930s. Available at SSRN 3739643 (2020).

23. Michney, T. M. How the City Survey’s Redlining Maps Were Made: A Closer Look at HOLC’s Mortgagee Rehabilitation Division. Journal of Planning History 15385132211013361 (2021).

24. Fishback, P., Rose, J., Snowden, K. A. & Storrs, T. New Evidence on Redlining by Federal Housing Programs in the 1930s. Journal of Urban Economics 141, 103462 (2024).

25. Gibbons, J. Linking U.S. government-sponsored redlining to early-stage white flight, 1940– 1950. Urban Geography 45, 1385–1406 (2024).

26. Markley, S. Federal ‘redlining’ maps: A critical reappraisal. Urban Studies 61, 195–213 (2024).

27. Swope, C. B., Hernández, D. & Cushing, L. J. The relationship of historical redlining with present-day neighborhood environmental and health outcomes: a scoping review and conceptual model. Journal of Urban Health 99, 959–983 (2022).

28. Senathirajah, M., Dankwa-Mullan, I., Pickens, G., Benevent, R. & Spurlock, B. A hospital social needs index would help hospitals collaborate to address social needs and health equity. Health Affairs Forefront (2022).

29. Srivastava, T., Schmidt, H., Sadecki, E. & Kornides, M. L. Disadvantage indices deployed to promote equitable allocation of COVID-19 vaccines in the US: a scoping review of differences and similarities in design. in JAMA health forum vol. 3 e214501–e214501 (American Medical Association, 2022).

30. Dasgupta, S. et al. Association between social vulnerability and a county’s risk for becoming a COVID-19 hotspot—United States, June 1–July 25, 2020. Morbidity and Mortality Weekly Report 69, 1535 (2020).

31. Price-Haywood, E. G., Burton, J., Fort, D. & Seoane, L. Hospitalization and mortality among black patients and white patients with Covid-19. New England Journal of Medicine 382, 2534–2543 (2020).

32. Hooper, M. W., Nápoles, A. M. & Pérez-Stable, E. J. COVID-19 and racial/ethnic disparities. JAMA 323, 2466–2467 (2020).

33. Tai, D. B. G., Shah, A., Doubeni, C. A., Sia, I. G. & Wieland, M. L. The Disproportionate Impact of COVID-19 on Racial and Ethnic Minorities in the United States. Clin Infect Dis (2020) doi:10.1093/cid/ciaa815.

34. Muñoz-Price, L. S. et al. Racial disparities in incidence and outcomes among patients with COVID-19. JAMA network open 3, e2021892–e2021892 (2020).

35. Azar, K. M. et al. Disparities in outcomes among COVID-19 patients in a large health care system in California: Study estimates the COVID-19 infection fatality rate at the US county level. Health Affairs 39, 1253–1262 (2020).

36. Reitsma, M. B. et al. Racial/Ethnic Disparities In COVID-19 Exposure Risk, Testing, And Cases At The Subcounty Level In California: Study examines racial/ethnic disparities in COVID-19 risk, testing, and cases. Health Affairs 40, 870–878 (2021).

37. Wrigley-Field, E., Berry, K. M., Stokes, A. C. & Leider, J. P. COVID-19 Vaccination and Racial/Ethnic Inequities in Mortality at Midlife in Minnesota. American Journal of Preventive Medicine (2022).

38. Wong, M. S., Brown, A. F. & Washington, D. L. Inclusion of Race and Ethnicity With Neighborhood Socioeconomic Deprivation When Assessing COVID-19 Hospitalization Risk Among California Veterans Health Administration Users. JAMA Network Open 6, e231471–e231471 (2023).

39. Bakhsh, Y., Readhead, A., Flood, J. & Barry, P. Association of Area-Based Socioeconomic Measures with Tuberculosis Incidence in California. Journal of Immigrant and Minority Health 1–10 (2022).

40. Maizlish, N. et al. California healthy places index: frames matter. Public Health Reports 134, 354–362 (2019).

41. Delaney, T. et al. Healthy Places Index (HPI 2.0). https://phasocal.org/wp-content/uploads/2018/04/HPI2Documentation2018-04-04-FINAL.pdf (2018).

42. Bodenreider, C. et al. Healthy Places Index (3.0). https://assets.website-files.com/613a633a3add5db901277f96/63320a9e98493bbdcc03d509_HPI3TechnicalReport2022-09-20.pdf (2022).

43. Nelson, R. K., Winling, L., Marciano, R., Connolly, N., &, et al. Mapping Inequality: Redlining in New Deal America, version 3.0. https://dsl.richmond.edu/panorama/redlining/data (2023).

44. Centers for Disease Control and Prevention/ Agency for Toxic Substances and Disease Registry/ Geospatial Research, Analysis, and Services Program. CDC/ATSDR Social Vulnerability Index 2020 Database California. https://www.atsdr.cdc.gov/placeandhealth/svi/data_documentation_download.html.

45. Krieger, N., Chen, J. T. & Waterman, P. D. The Public Health Disparities Geocoding Project. https://www.hsph.harvard.edu/thegeocodingproject/ (2022).

46. Unversity of Wisconsin School of Medicine and Public Health. Area Deprivation Index. https://www.neighborhoodatlas.medicine.wisc.edu/ (2022).

47. Shete, P., Vargo, J., Chen, A. & Bibbins-Domingo, K. Equity metrics: Toward a more effective and inclusive pandemic response. Health Affairs Blog 2, 21 (2021).

48. California Department of Public Health. Blueprint For a Safer Economy: Equity Focus. https://www.cdph.ca.gov/Programs/CID/DCDC/Pages/COVID-19/CaliforniaHealthEquityMetric.aspx (2021).

49. Hoover, C. M. et al. California’s COVID-19 Vaccine Equity Policy: Cases, Hospitalizations, And Deaths Averted In Affected Communities: Study examines California’s COVID-19 vaccine equity policy. Health Affairs 43, 632–640 (2024).

50. GitHub Repository. https://github.com/Cesariddle/Index-Comparison-Analysis.

51. Wickham, H. et al. Welcome to the Tidyverse. Journal of open source software 4, 1686 (2019).

52. Brunson, J. C. & Read, Q. D. Package ‘ggalluvial’. (2019).

53. Public Health Alliance of Southern California. California Healthy Places Index. https://www.healthyplacesindex.org/ (2022).

54. Singh, G. K. Area deprivation and widening inequalities in US mortality, 1969–1998. American journal of public health 93, 1137–1143 (2003).

55. Maroko, A. R., et al. Peer Reviewed: Integrating Social Determinants of Health With Treatment and Prevention: A New Tool to Assess Local Area Deprivation. Preventing chronic disease 13, (2016).

56. U.S. Census Bureau. 2015-2019 ACS 5-year Estimates. (2020).

57. White, L. A., McCorvie, R., Crow, D., Jain, S. & León, T. M. Assessing the accuracy of California county level COVID-19 hospitalization forecasts to inform public policy decision making. BMC Public Health 23, 782 (2023).

58. Havard, S. et al. A small-area index of socioeconomic deprivation to capture health inequalities in France. Social science & medicine 67, 2007–2016 (2008).

59. Kwan, A. T. et al. The integration of health equity into policy to reduce disparities: Lessons from California during the COVID-19 pandemic. PloS one 20, e0316517 (2025).

60. Moss, J. L., Johnson, N. J., Yu, M., Altekruse, S. F. & Cronin, K. A. Comparisons of individual- and area-level socioeconomic status as proxies for individual-level measures: evidence from the Mortality Disparities in American Communities study. Popul Health Metrics 19, 1 (2021).

61. Cho, W. K. T. & Hwang, D. G. Differential effects of race/ethnicity and social vulnerability on COVID-19 positivity, hospitalization, and death in the San Francisco Bay area. Journal of Racial and Ethnic Health Disparities 10, 834–843 (2023).

62. California Secretary of State. Proposition 209: Text of Proposed Law. https://vigarchive.sos.ca.gov/1996/general/pamphlet/209text.htm.

