## Supplement for "A Comparative Analysis of Area-Based Socioeconomic Measures: Implications for Future Equity-focused Public Health Response"

### Supplement A1. ABSM constituents

| **ABSM**, Variable Domains, Variables | | **Data Source†, Year** |
|---|---|---|
| **California Healthy Places Index v3.0 (HPI)** | | **Public Health Alliance of Southern California, 2020** |
| **1** | **Economic** | |
| | Percent of the population with an income exceeding 200% of federal poverty level | ACS, 2015-2019 |
| | Percentage of population aged 25-64 who are employed | ACS, 2015-2019 |
| | Median Household Income | - |
| | Per capita income | ACS, 2015-2019 |
| **2** | **Education** | |
| | Percentage of population over age 25 with a bachelor's education or higher | ACS, 2015-2019 |
| | Percentage of 15–17-year-olds enrolled in school | ACS, 2015-2019 |
| | Percentage of 3- and 4-year-olds enrolled in pre-school | ACS, 2015-2019 |
| **3** | **Social** | |
| | Percentage of registered voters voting in the 2020 general election | UC Berkeley, 2020 |
| | Percentage of family households with children under 18 with two parents | - |
| | Percent of the population responding to the 2020 census (short form) | Census, 2020 |
| **4** | **Transportation** | |
| | Percentage of households with access to an automobile | ACS, 2015-2019 |
| | Percentage of workers (16 years and older) commuting by walking, cycling, or transit (excluding working from home) | ACS, 2015-2019 |
| **5** | **Healthcare Access** | |
| | Percentage of adults aged 18 to 64 years currently insured | ACS, 2015-2019 |
| **6** | **Neighborhood** | |
| | Percentage of the population living within ½ -mile of a park, beach, or open space greater than 1 acre | GreenInfo, 2012 |
| | Population-weighted percentage of the census tract area with tree canopy | NLCD, 2011 |
| | Percentage of the population residing within ¼ mile of an off-site sales alcohol outlet | - |
| | Percentage of the urban and small-town population residing less than 1/2 mile from a supermarket/large grocery store, and the percent of the rural population living less than 1 miles from a supermarket/large grocery store | - |
| | Combined employment density for retail, entertainment, supermarkets, and educational uses (jobs/acre) | LODES, 2014-2018 |
| **7** | **Housing** | |
| | Percentage of occupied housing units occupied by property owners | ACS, 2015-2019 |
| | Percent of households with complete kitchen facilities and plumbing | CHAS, 2014-2018 |
| | Percentage of low-income homeowners paying more than 50% of income on housing | CHAS, 2014-2018 |
| | Percentage of low-income renter households paying more than 50% of income on housing | CHAS, 2013-2017 |
| | Percentage of households with less or equal to 1 occupant per room | ACS, 2015-2019 |
| **8** | **Clean Environment** | |
| | Annual average spatial distribution of gridded diesel PM emissions from on-road and non-road sources 2016 (tons/year). | CalEPA, 2016 |
| | CalEnviroScreen 4.0 drinking water contaminant index for selected contaminants | CalEPA, 2011-2019 |
| | Mean of summer months (May-October) of the daily maximum 8-hour ozone concentration (ppm), averaged over three years (2017 to 2019) | CalEPA, 2017-2019 |

| | |
|---|---|
| Annual mean concentration of PM2.5 (μg/m3) over three years (2015 to 2017). | CalEPA, 2015-2017 |
| **Area Deprivation Index (ADI)** | **Kind and Buckingham, 2018; University of Wisconsin School of Medicine and Public Health, 2020.** |
| **Education** | |
| Percent of the block group's population aged ≥ 25 years with < 9 years of education | ACS |
| Percent aged ≥ 25 years with greater than or equal to a high school diploma | ACS |
| Percent of employed persons ≥16 years of age in white-collar occupations | ACS |
| **Income/Employment** | |
| Median family income | ACS |
| Income disparity | ACS |
| Percent of families below the poverty level | ACS |
| Percent of population below 150% of the poverty threshold | ACS |
| Percent of civilian labor force population ≥ 16 years of age unemployed (unemployment rate) | ACS |
| **Housing** | |
| Median home value | ACS |
| Median gross rent | ACS |
| Median monthly mortgage | ACS |
| Percent owner-occupied housing units (home ownership rate) | ACS |
| Percent of occupied housing units without complete plumbing (log) | ACS |
| **Household Characteristics** | |
| Percent of single-parent households with children < 18 years of age | ACS |
| Percent of occupied housing units without a motor vehicle | ACS |
| Percent of occupied housing units without a telephone | ACS |
| Percent of occupied housing units without complete plumbing (log) | ACS |
| Percent of occupied housing units with more than one person per room (crowding) | ACS |
| **Social Vulnerability Index (SVI)** | **CDC/ATSDR, 2020** |
| **Socioeconomic Status** | |
| Below 150% Poverty | ACS: S1701_C01_040E |
| Unemployed | ACS: DP03_0005E |
| Housing Cost Burden | ACS: S2503_C01_028E + S2503_C01_032E + S2503_C01_036E + S2503_C01_040E |
| No High School Diploma | ACS: B06009_002E |
| No Health Insurance | ACS: S2701_C04_001E |
| **Household Characteristics** | |
| Aged 65 & Older | ACS: S0101_C01_030E |
| Aged 17 & Younger | ACS: B09001_001E |
| Civilian with a Disability | ACS: DP02_0072E |
| Single-Parent Households with Children under 18 | ACS: B11012_010E + B11012_015E |
| English Language Proficiency among Persons (age 5+) who speak English "less than well" | ACS: B16005_007E + B16005_008E + B16005_012E + B16005_013E + B16005_017E + B16005_018E + B16005_022E + B16005_023E + B16005_029E + B16005_030E + |

| | | |
|---|---|---|
| | | B16005_034E + B16005_035E + B16005_039E + B16005_040E + B16005_044E + B16005_045E |
| | **Racial & Ethnic Minority Status** | |
| | Hispanic or Latino (of any race); Black and African American, Not Hispanic or Latino; American Indian and Alaska Native, Not Hispanic or Latino; Asian, Not Hispanic or Latino; Native Hawaiian and Other Pacific Islander, Not Hispanic or Latino; Two or More Races, Not Hispanic or Latino; Other Races, Not Hispanic or Latino 4 | ACS: DP05_0071E + DP05_0078E + DP05_0079E + DP05_0080E + DP05_0081E + DP05_0082E + DP05_0083E |
| | **Housing Type & Transportation** | |
| | Multi-Unit Structures (Housing in structures with 10 or more units) | ACS: DP04_0012E + DP04_0013E |
| | Mobile Homes | ACS: DP04_0014E |
| | Crowding (more people than rooms) | ACS: DP04_0078E + DP04_0079E |
| | No Vehicle | ACS: DP04_0058E |
| | Group Quarters (Persons in group quarters) | ACS: B26001_001E |
| **Index of Concentration at the Extremes (ICE)** | | **Krieger, Chen, Waterman, 2020** |
| | Total Population | ACS: B01003_001E |
| | White Non-Hispanic Population | ACS: B01001H_001E |
| | % of persons below poverty | ACS: B17001_002E / B17001_001E |
| | Index of Concentration at the Extremes (high-income white households versus low-income black households) | ACS: ((B19001A_014E + B19001A_015E + B19001A_016E + B19001A_017E) – (B19001B_002E + B19001B_003E + B19001B_004E + B19001B_005E)) / B19001_001E |
| | Index of Concentration at the Extremes (high-income white non-Hispanic households versus low-income people of color households) | ACS: (B19001H_014E + B19001H_015E + B19001H_016E + B19001H_017E) – [(B19001_002E + B19001_003E + B19001_004E + B19001_005E) – (B19001H_002E + B19001H_003E + B19001H_004E + B19001H_005E)]/ B19001_001E |
| | % crowding (>1 person per room) | ACS: (B25014_005E + B25014_006E + B25014_007E + B25014_011E + B25014_012E + B25014_013E) / B25014_001E |
| | % population of color (not White Non-Hispanic) | ACS: (B01003_001E -B01001H_001E) / B01003_001E |
| **Home Owner's Loan Corporation (HOLC) "Redlining" Scores** | | **Fishback, LaVoice, 2021; Nelson et al. 2023.** |
| | HOLC residential security grades:<br>Areas with grades A ("best") and B ("still desirable") were seen as "more desirable and less risky" for mortgage lending.<br>Areas with grades C ("definitely declining") and D ("hazardous") were seen as "less desirable and riskier." | |

**Notes.**
HPI source: https://assets.website-files.com/613a633a3add5db901277f96/63320a9e98493bbdcc03d509_HPI3TechnicalReport2022-09-20.pdf
ADI source: https://www.neighborhoodatlas.medicine.wisc.edu/login
SVI source: https://www.atsdr.cdc.gov/placeandhealth/svi/img/pdf/Flanagan_2011_SVIforDisasterManagement-508.pdf
ICE source: https://www.hsph.harvard.edu/thegeocodingproject/covid-19-resources/
HOLC grades source: https://www.openicpsr.org/openicpsr/project/141121/version/V2/view

**Supplement A2. Maps of California census tracts by ABSM deciles**

a. HPI v3.0

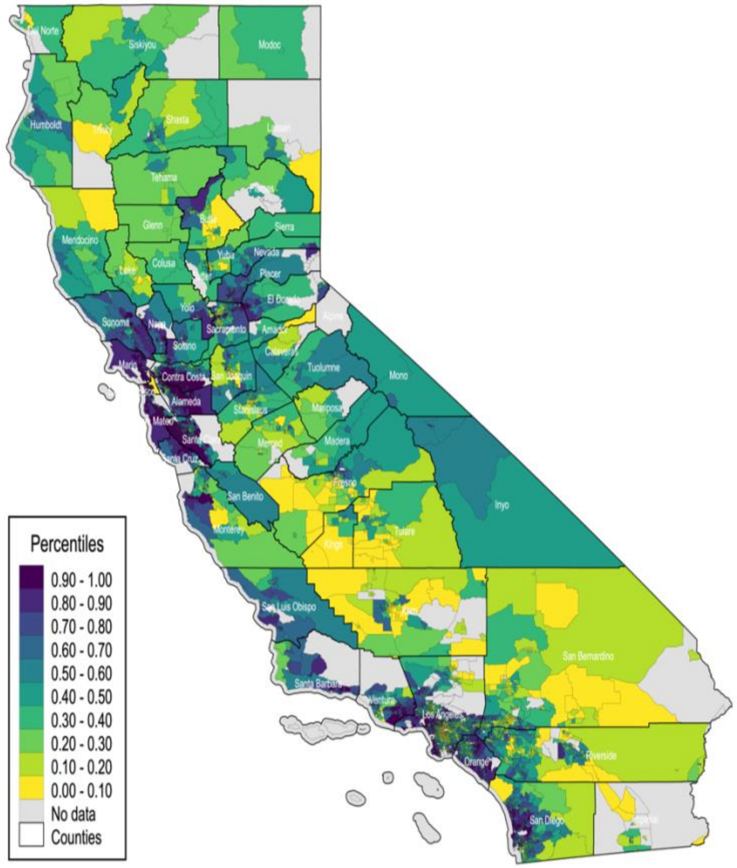


b. ADI 2019 (transformed)

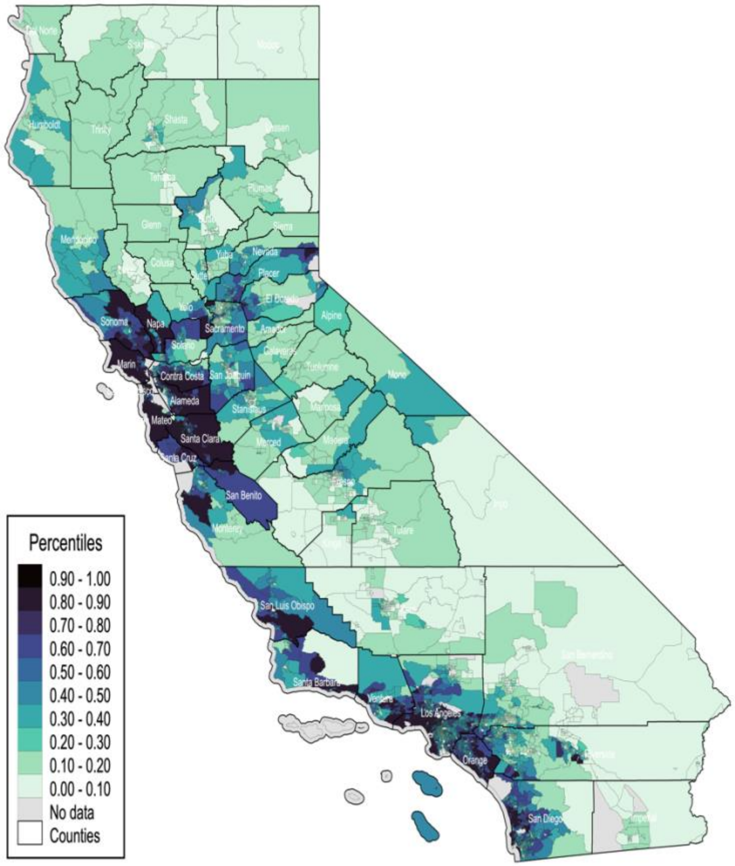

c. SVI 2020 (transformed)

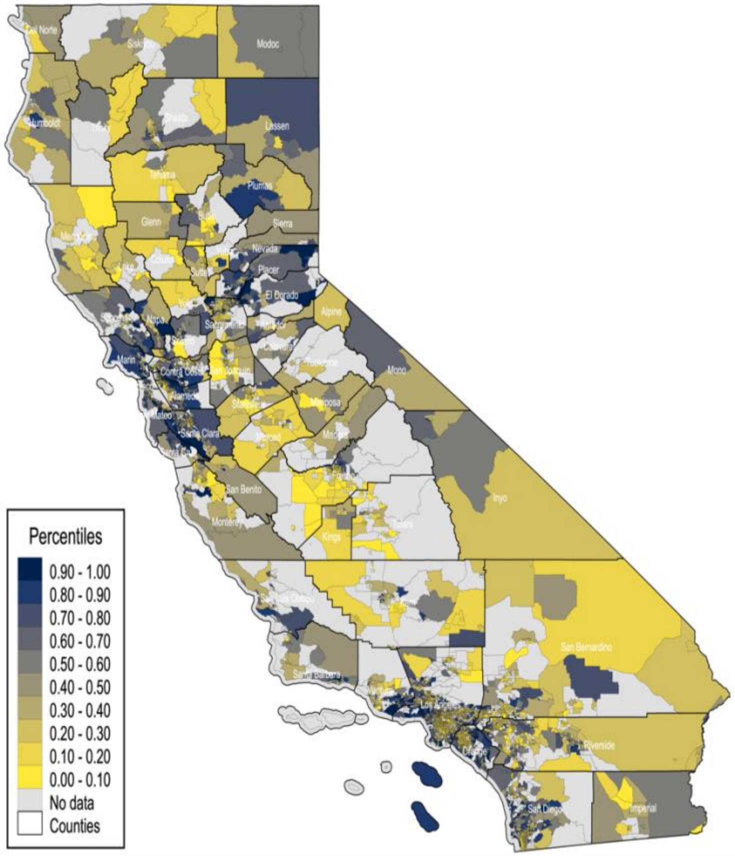


d. ICE 2020 (transformed)

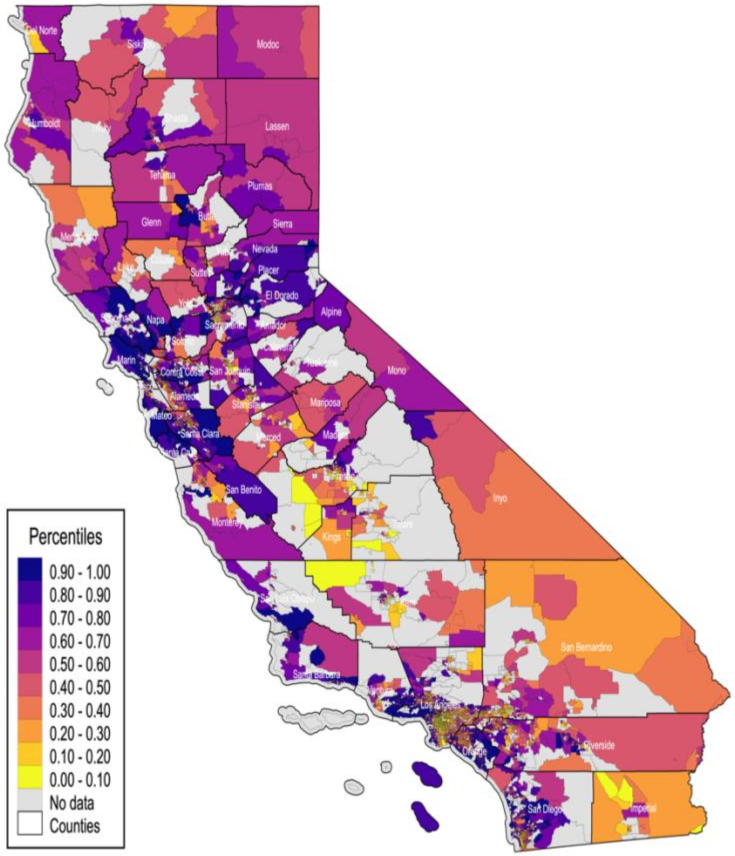


**Note:** HPI is California Healthy Places Index, version 3.0. ADI is Area Deprivation Index. SVI is Social Vulnerability Index. ICE is Index of Concentration at the Extremes. ADI, SVI, and ICE are transformed as described in Table 1. Census tracts are based on the boundaries delineated in the 2010 US Census.

### Supplement A3. Maps of California census tracts by CDPH regions and ABSM deciles

a. Northern California Region

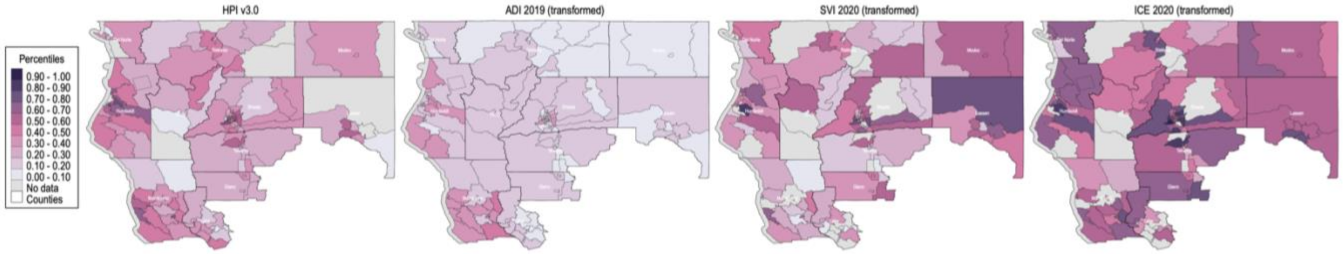


b. Bay Area Region

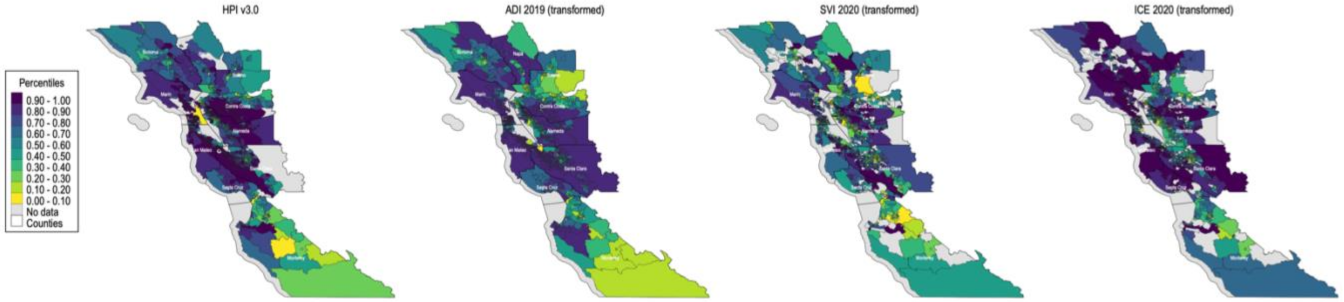


c. Greater Sacramento Region

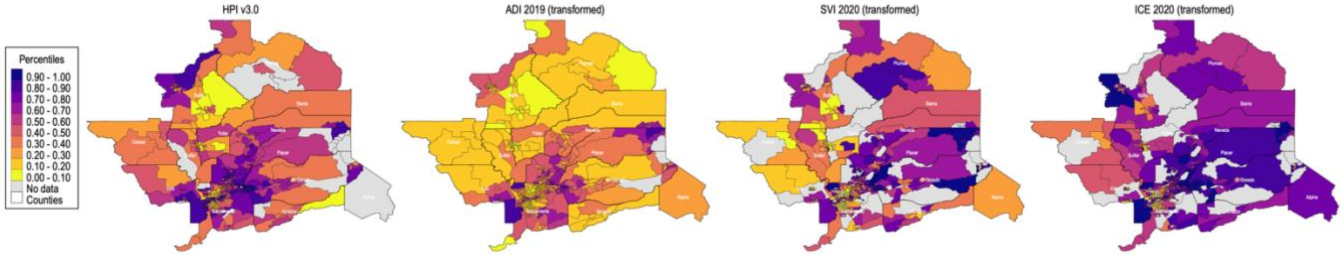


d. San Joaquin Valley Region

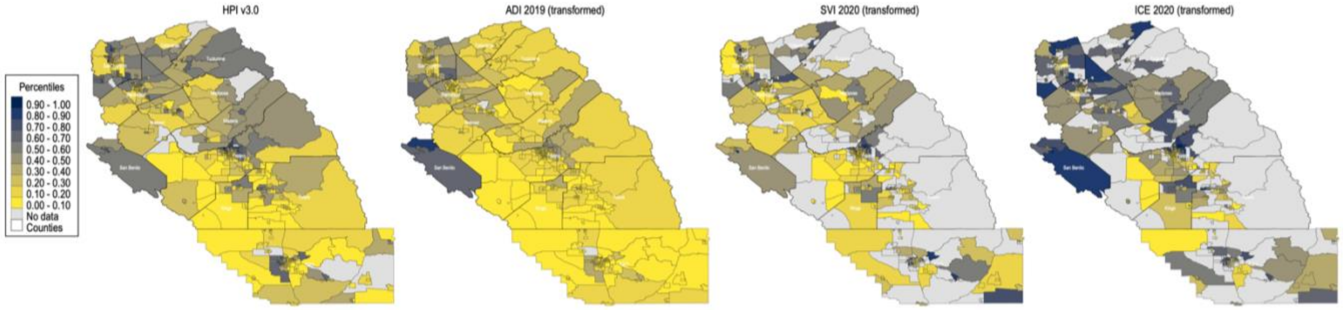


e. Southern California Region

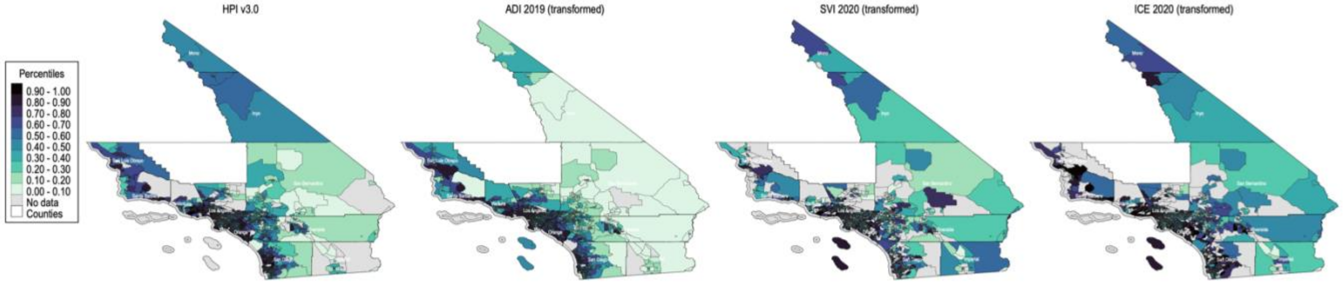


**Notes:** HPI is California Healthy Places Index, version 3.0. ADI is Area Deprivation Index. SVI is Social Vulnerability Index. ICE is Index of Concentration at the Extremes. ADI, SVI, and ICE are transformed as described in Table 1. Census tracts are based on the boundaries delineated in the 2010 US Census.

**Supplement A4. Descriptive statistics: HPI variable domains (policy action areas) and corresponding constituent determinants for census tracts in California pooled, by county size, and city sample**

| Policy Action Area | | A. All Counties in California (N=8,057 census tracts) | | | B. Large County Sample (N=7,839 census tracts) | | | C. Small County Sample (N=218 census tracts) | | | D. City Sample (N=1,120 census tracts) | | |
|---|---|---|---|---|---|---|---|---|---|---|---|---|---|
| | *Constituent* | *N* | *Mean or Prop.* | *SD* | *N* | *Mean or Prop.* | *SD* | *N* | *Mean or Prop.* | *SD* | *N* | *Mean or Prop.* | *SD* |
| **Economic** | | | | | | | | | | | | | |
| *1* | *Prop. of population with income exceeding 200% of federal poverty level* | 7,790 | 0.69 | 0.18 | 7,591 | 0.69 | 0.18 | 199 | 0.64 | 0.13 | 1,117 | 0.62 | 0.19 |
| *2* | *Prop. of population aged 25-64 employed* | 7,790 | 0.73 | 0.08 | 7,591 | 0.73 | 0.08 | 199 | 0.64 | 0.09 | 1,117 | 0.75 | 0.07 |
| *3* | *Per capita income* | 7,790 | 38,067.94 | 22,284.93 | 7,591 | 38,273.70 | 22,497.58 | 199 | 30,218.70 | 8,398.90 | 1,117 | 38,413.97 | 26,853.34 |
| **Education** | | | | | | | | | | | | | |
| 4 | *Prop. of population >25yrs with a bachelor's education or higher* | 7,790 | 0.33 | 0.21 | 7,591 | 0.34 | 0.21 | 199 | 0.21 | 0.10 | 1,117 | 0.36 | 0.25 |
| 5 | *Prop. of 15-17-year-olds enrolled in school* | 7,790 | 0.98 | 0.07 | 7,591 | 0.98 | 0.07 | 199 | 0.97 | 0.07 | 1,117 | 0.97 | 0.08 |
| 6 | *Prop. of 3-4-year-olds enrolled in pre-school* | 7,790 | 0.53 | 0.28 | 7,591 | 0.53 | 0.28 | 199 | 0.48 | 0.29 | 1,117 | 0.60 | 0.28 |
| **Social** | | | | | | | | | | | | | |
| *7* | *Prop. of 2020 decennial households who completed census forms online, by mail, or by phone* | 7,790 | 0.70 | 0.11 | 7,591 | 0.71 | 0.11 | 199 | 0.59 | 0.14 | 1,117 | 0.65 | 0.10 |
| *8* | *Prop. of registered voters voting in 2020 general election* | 7,790 | 0.77 | 0.10 | 7,591 | 0.77 | 0.10 | 199 | 0.80 | 0.07 | 1,117 | 0.72 | 0.11 |
| **Transportation** | | | | | | | | | | | | | |
| *9* | *Prop. of households with access to an automobile* | 7,790 | 0.93 | 0.08 | 7,591 | 0.93 | 0.08 | 199 | 0.95 | 0.04 | 1,117 | 0.87 | 0.09 |
| *10* | *Prop. of workers (16 years and older) commuting by walking, cycling, or transit* | 7,790 | 0.09 | 0.12 | 7,591 | 0.09 | 0.12 | 199 | 0.05 | 0.05 | 1,117 | 0.19 | 0.16 |
| **Healthcare Access** | | | | | | | | | | | | | |
| *11* | *Prop. of adults 18-64yrs currently insured* | 7,790 | 0.89 | 0.08 | 7,591 | 0.89 | 0.08 | 199 | 0.89 | 0.05 | 1,117 | 0.85 | 0.10 |
| **Neighborhood** | | | | | | | | | | | | | |
| *12* | *Prop. of population living within ½ -mile of a park, beach, or open space >1 acre* | 7,790 | 0.77 | 0.31 | 7,591 | 0.77 | 0.31 | 199 | 0.52 | 0.32 | 1,117 | 0.79 | 0.32 |

| | | | | | | | | | | | | |
|---|---|---|---|---|---|---|---|---|---|---|---|---|
| 13 | *Population-weighted proportion of census tract area with tree canopy* | 7,790 | 0.08 | 0.09 | 7,591 | 0.08 | 0.08 | 199 | 0.23 | 0.18 | 1,117 | 0.06 | 0.05 |
| 14 | *Employment density for retail, entertainment, supermarkets & education (jobs/acre)* | 7,790 | 6.96 | 21.84 | 7,591 | 7.11 | 22.10 | 199 | 1.29 | 2.67 | 1,117 | 10.88 | 21.65 |
| **Housing** | | | | | | | | | | | | | |
| 15 | *Prop. of occupied housing units occupied by property owners* | 7,790 | 0.55 | 0.24 | 7,591 | 0.55 | 0.24 | 199 | 0.66 | 0.16 | 1,117 | 0.35 | 0.21 |
| 16 | *Prop. of households with complete kitchen facilities and plumbing* | 7,790 | 0.99 | 0.02 | 7,591 | 0.99 | 0.02 | 199 | 0.99 | 0.02 | 1,117 | 0.98 | 0.02 |
| 17 | *Prop. of low-income homeowners paying >50% of income on housing* | 7,790 | 0.12 | 0.08 | 7,591 | 0.12 | 0.08 | 199 | 0.10 | 0.04 | 1,117 | 0.16 | 0.11 |
| 18 | *Prop. of low-income renter households paying >50% of income on housing* | 7,790 | 0.25 | 0.11 | 7,591 | 0.25 | 0.11 | 199 | 0.23 | 0.10 | 1,117 | 0.28 | 0.10 |
| 19 | *Prop. of households with ≤1 occupant per room* | 7,790 | 0.91 | 0.09 | 7,591 | 0.91 | 0.09 | 199 | 0.96 | 0.03 | 1,117 | 0.86 | 0.11 |
| **Clean Environment** | | | | | | | | | | | | | |
| 20 | *Annual average spatial distribution of gridded diesel PM emissions from on-road & non-road sources (tons/year)* | 7,790 | 0.22 | 0.30 | 7,591 | 0.23 | 0.31 | 199 | 0.05 | 0.07 | 1,117 | 0.34 | 0.28 |
| 21 | *CalEnviroScreen 4.0 drinking water contaminant index for selected contaminants* | 7,790 | 477.36 | 218.19 | 7,591 | 478.86 | 219.39 | 199 | 420.38 | 156.13 | 1,117 | 521.79 | 246.54 |
| 22 | *Mean of summer months (May-Oct) of daily max 8-hour ozone concentration (ppm) averaged over 2017, 2018, 2019* | 7,790 | 0.05 | 0.01 | 7,591 | 0.05 | 0.01 | 199 | 0.05 | 0.01 | 1,117 | 0.04 | 0.01 |
| 23 | *Annual mean concentration of PM2.5 (µg/m3) over three years (2015-17)* | 7,790 | 10.18 | 2.15 | 7,591 | 10.27 | 2.07 | 199 | 6.52 | 1.97 | 1,117 | 11.17 | 1.30 |

**Notes:** Large counties are California counties with population >106,000; small counties have populations with 106,000 or fewer. The city sample includes any tract with a HOLC score. Prop is proportion; SD is standard deviation. HPI is California Healthy Places Index, version 3.0. All economic, education, transportation, and healthcare access measures are from ACS 2015-2019 5-year estimates. Registered voters is from UC Berkeley 2020. Census response is from Decennial census 2020. Neighborhood measures are from Greeninfo (2012), NLCD (2011), and USEPA (2010), respectively. Housing measures are from ACS 2015-2019 5-year estimates or CHAS 2010-2014. Clean environment variables are from CalEPA (2016), CalEPA (2011-2019), CalEPA (2017-2019), and CalEPA (2015-2017), respectively. HPI v3 source: https://assets.website-files.com/613a633a3add5db901277f96/63320a9e98493bbdcc03d509_HPI3TechnicalReport2022-09-20.pdf. Census tracts are based on the boundaries delineated in the 2010 US Census.

### Supplement A5. 2x2 frequency tables of California census tracts across statewide ABSM quartiles

| | ADI 2019 (transformed) | | | | | |
|---|---|---|---|---|---|---|
| | Q1 | Q2 | Q3 | Q4 | Missing | Total |
| | *N* | *N* | *N* | *N* | *N* | *N* |
| HPI v3.0 | *(Row %)* | *(Row %)* | *(Row %)* | *(Row %)* | *(Row %)* | *(Col %)* |
| Q1 | 1,223 | 510 | 190 | 20 | 5 | 1,948 |
| | 62.8% | 26.2% | 9.8% | 1.0% | 0.3% | 24.2% |
| Q2 | 702 | 746 | 455 | 43 | 1 | 1,947 |
| | 36.1% | 38.3% | 23.4% | 2.2% | 0.1% | 24.2% |
| Q3 | 197 | 547 | 959 | 244 | 1 | 1,948 |
| | 10.1% | 28.1% | 49.2% | 12.5% | 0.1% | 24.2% |
| Q4 | 6 | 83 | 627 | 1,231 | 0 | 1,947 |
| | 0.3% | 4.3% | 32.2% | 63.2% | 0.0% | 24.2% |
| Missing | 50 | 23 | 35 | 27 | 132 | 267 |
| | 18.7% | 8.6% | 13.1% | 10.1% | 49.4% | 3.3% |
| Total | 2,178 | 1,909 | 2,266 | 1,565 | 139 | 8,057 |
| | 27.0% | 23.7% | 28.1% | 19.4% | 1.7% | 100.0% |

| | SVI 2020 (transformed) | | | | | |
|---|---|---|---|---|---|---|
| | Q1 | Q2 | Q3 | Q4 | Missing | Total |
| | *N* | *N* | *N* | *N* | *N* | *N* |
| HPI v3.0 | *(Row %)* | *(Row %)* | *(Row %)* | *(Row %)* | *(Row %)* | *(Col %)* |
| Q1 | 1,217 | 386 | 47 | 16 | 282 | 1,948 |
| | 62.5% | 19.8% | 2.4% | 0.8% | 14.5% | 24.2% |
| Q2 | 443 | 814 | 366 | 41 | 283 | 1,947 |
| | 22.8% | 41.8% | 18.8% | 2.1% | 14.5% | 24.2% |
| Q3 | 34 | 417 | 802 | 366 | 329 | 1,948 |
| | 1.7% | 21.4% | 41.2% | 18.8% | 16.9% | 24.2% |
| Q4 | 2 | 57 | 451 | 1,216 | 221 | 1,947 |
| | 0.1% | 2.9% | 23.2% | 62.5% | 11.4% | 24.2% |
| Missing | 8 | 27 | 36 | 63 | 133 | 267 |
| | 3.0% | 10.1% | 13.5% | 23.6% | 49.8% | 3.3% |
| Total | 1,704 | 1,701 | 1,702 | 1,702 | 1248 | 8,057 |
| | 21.1% | 21.1% | 21.1% | 21.1% | 15.5% | 100.0% |

| | ICE 2020 (transformed) | | | | | |
|---|---|---|---|---|---|---|
| | Q1 | Q2 | Q3 | Q4 | Missing | Total |
| | *N* | *N* | *N* | *N* | *N* | *N* |
| HPI v3.0 | *(Row %)* | *(Row %)* | *(Row %)* | *(Row %)* | *(Row %)* | *(Col %)* |
| Q1 | 1,252 | 350 | 63 | 1 | 282 | 1,948 |
| | 64.3% | 18.0% | 3.2% | 0.1% | 14.5% | 24.2% |
| Q2 | 377 | 827 | 414 | 46 | 283 | 1,947 |
| | 19.4% | 42.5% | 21.3% | 2.4% | 14.5% | 24.2% |
| Q3 | 39 | 405 | 794 | 381 | 329 | 1,948 |
| | 2.0% | 20.8% | 40.8% | 19.6% | 16.9% | 24.2% |
| Q4 | 2 | 90 | 404 | 1,230 | 221 | 1,947 |
| | 0.1% | 4.6% | 20.7% | 63.2% | 11.4% | 24.2% |
| Missing | 33 | 31 | 28 | 44 | 131 | 267 |
| | 12.4% | 11.6% | 10.5% | 16.5% | 49.1% | 3.3% |
| Total | 1,703 | 1,703 | 1,703 | 1,702 | 1246 | 8,057 |
| | 21.1% | 21.1% | 21.1% | 21.1% | 15.5% | 100.0% |

| | SVI 2020 (transformed) | | | | | |
|---|---|---|---|---|---|---|
| | Q1 | Q2 | Q3 | Q4 | Missing | Total |
| | *N* | *N* | *N* | *N* | *N* | *N* |
| ADI 2019 (transformed) | *(Row %)* | *(Row %)* | *(Row %)* | *(Row %)* | *(Row %)* | *(Col %)* |
| Q1 | 921 | 568 | 269 | 57 | 363 | 2,178 |
| | 42.3% | 26.1% | 12.4% | 2.6% | 16.7% | 27.0% |
| Q2 | 496 | 567 | 391 | 180 | 275 | 1,909 |
| | 26.0% | 29.7% | 20.5% | 9.4% | 14.4% | 23.7% |
| Q3 | 259 | 473 | 656 | 553 | 325 | 2,266 |
| | 11.4% | 20.9% | 28.9% | 24.4% | 14.3% | 28.1% |
| Q4 | 26 | 89 | 375 | 886 | 189 | 1,565 |
| | 1.7% | 5.7% | 24.0% | 56.6% | 12.1% | 19.4% |
| Missing | 2 | 4 | 11 | 26 | 96 | 139 |
| | 1.4% | 2.9% | 7.9% | 18.7% | 69.1% | 1.7% |
| Total | 1,704 | 1,701 | 1,702 | 1,702 | 1248 | 8,057 |
| | 21.1% | 21.1% | 21.1% | 21.1% | 15.5% | 100.0% |

| | ICE 2020 (transformed) | | | | | |
|---|---|---|---|---|---|---|
| | Q1 | Q2 | Q3 | Q4 | Missing | Total |
| | *N* | *N* | *N* | *N* | *N* | *N* |
| ADI 2019 (transformed) | *(Row %)* | *(Row %)* | *(Row %)* | *(Row %)* | *(Row %)* | *(Col %)* |
| Q1 | 823 | 600 | 352 | 40 | 363 | 2,178 |
| | 37.8% | 27.5% | 16.2% | 1.8% | 16.7% | 27.0% |
| Q2 | 560 | 484 | 419 | 171 | 275 | 1,909 |
| | 29.3% | 25.4% | 21.9% | 9.0% | 14.4% | 23.7% |
| Q3 | 285 | 474 | 609 | 574 | 324 | 2,266 |
| | 12.6% | 20.9% | 26.9% | 25.3% | 14.3% | 28.1% |
| Q4 | 22 | 131 | 316 | 907 | 189 | 1,565 |
| | 1.4% | 8.4% | 20.2% | 58.0% | 12.1% | 19.4% |
| Missing | 13 | 14 | 7 | 10 | 95 | 139 |
| | 9.4% | 10.1% | 5.0% | 7.2% | 68.3% | 1.7% |
| Total | 1,703 | 1,703 | 1,703 | 1,702 | 1246 | 8,057 |
| | 21.1% | 21.1% | 21.1% | 21.1% | 15.5% | 100.0% |

| | ICE 2020 (transformed) | | | | | |
|---|---|---|---|---|---|---|
| | Q1 | Q2 | Q3 | Q4 | Missing | Total |
| | *N* | *N* | *N* | *N* | *N* | *N* |
| SVI 2020 (transformed) | *(Row %)* | *(Row %)* | *(Row %)* | *(Row %)* | *(Row %)* | *(Col %)* |
| Q1 | 1,204 | 441 | 59 | 0 | 0 | 1,704 |
| | 70.7% | 25.9% | 3.5% | 0.0% | 0.0% | 21.1% |
| Q2 | 432 | 759 | 459 | 51 | 0 | 1,701 |
| | 25.4% | 44.6% | 27.0% | 3.0% | 0.0% | 21.1% |
| Q3 | 60 | 409 | 823 | 410 | 0 | 1,702 |
| | 3.5% | 24.0% | 48.4% | 24.1% | 0.0% | 21.1% |
| Q4 | 5 | 94 | 362 | 1,241 | 0 | 1,702 |
| | 0.3% | 5.5% | 21.3% | 72.9% | 0.0% | 21.1% |
| Missing | 2 | 0 | 0 | 0 | 1246 | 1,248 |
| | 0.2% | 0.0% | 0.0% | 0.0% | 99.8% | 15.5% |
| Total | 1,703 | 1,703 | 1,703 | 1,702 | 1246 | 8,057 |
| | 21.1% | 21.1% | 21.1% | 21.1% | 15.5% | 100.0% |

**Notes:** HPI is California Healthy Places Index, version 3.0. ADI is Area Deprivation Index. SVI is Social Vulnerability Index. ICE is Index of Concentration at the Extremes. ADI, SVI, and ICE are transformed as described in Table 1.

### Supplement A6. Maps of Los Angeles County census tracts by ABSM deciles

a. HPI v3.0

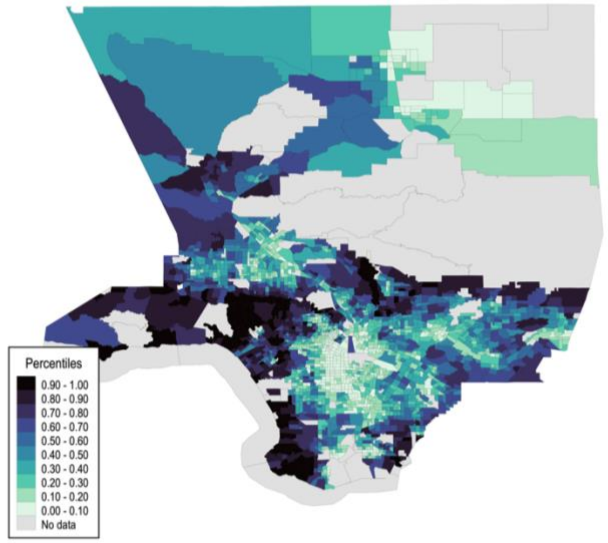


b. ADI 2019 (transformed)

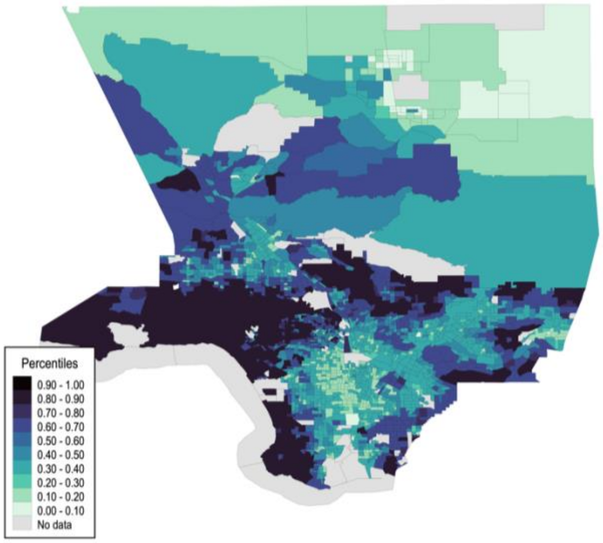


c. SVI 2020 (transformed)

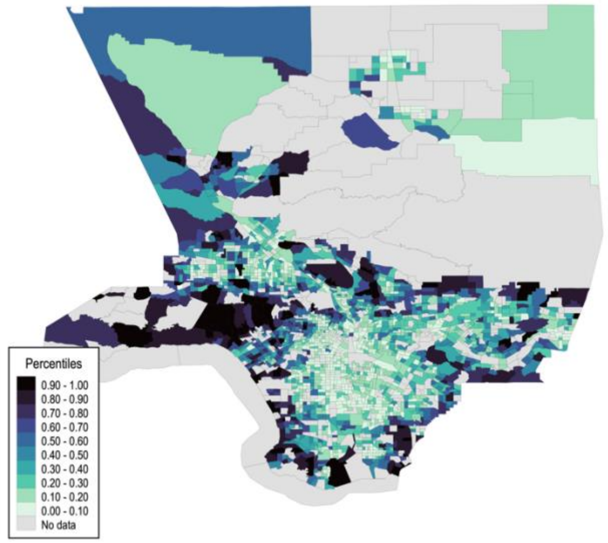


d. ICE 2020 (transformed)

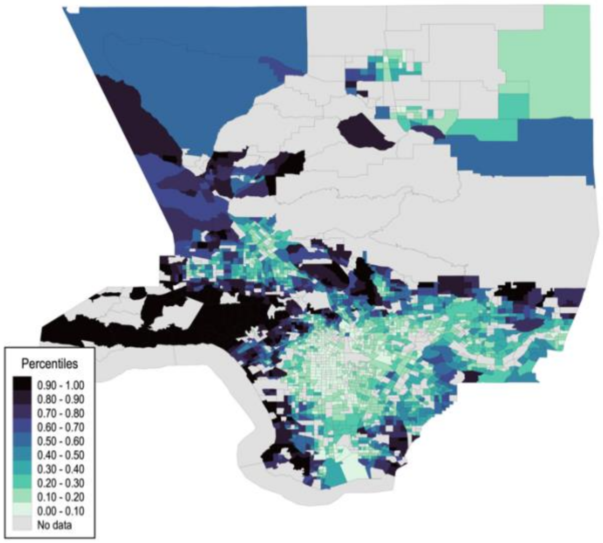


**Notes:** HPI is California Healthy Places Index, version 3.0. ADI is Area Deprivation Index. SVI is Social Vulnerability Index. ICE is Index of Concentration at the Extremes. ADI, SVI, and ICE are transformed as described in Table 1. Census tracts are based on the boundaries delineated in the 2010 US Census.

### Supplement A7. Correlations among statewide ABSM quartiles for tracts in large and small counties

a. Large Counties (County population is greater than 106,000)

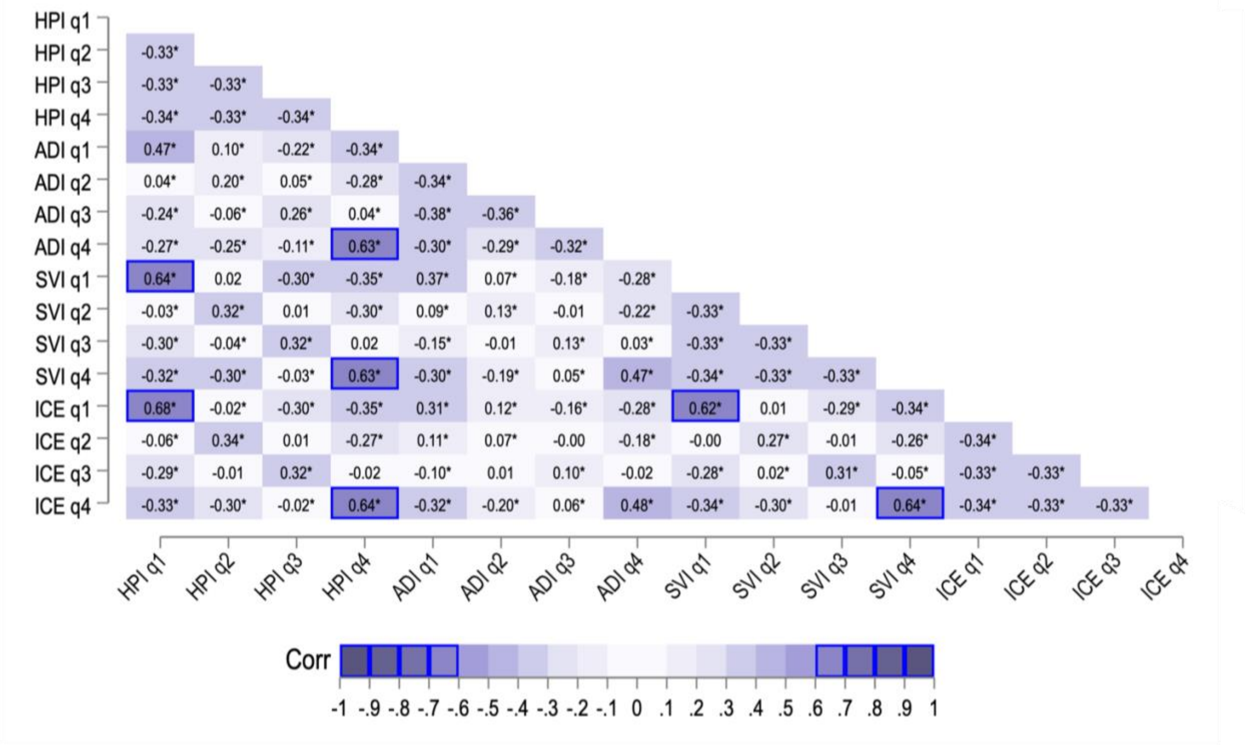


b. Small Counties (County population is less than or equal to 106,000)

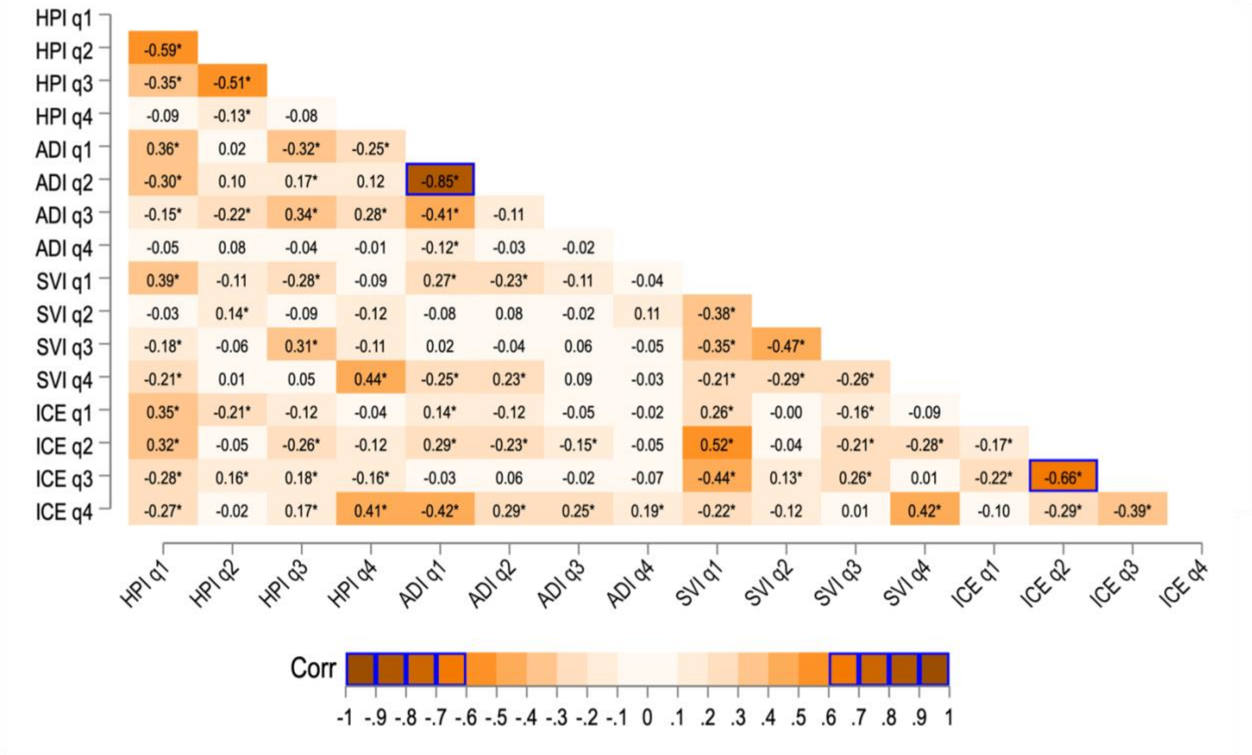


**Notes:** Values in each box are pairwise Pearson correlations (Corr) with significance designated as: * $p<0.1$. HPI is California Healthy Places Index, version 3.0. ADI is Area Deprivation Index. SVI is Social Vulnerability Index. ICE is Index of Concentration at the Extremes. ADI, SVI, and ICE are transformed as described in Table 1. Labels "q1" to "q4" refer to quartile 1 to quartile 4 for corresponding measure, with HPI q1 (2,3,4) referring to HPI quartile 1 (2,3,4). Blue boxes represent a correlation value greater than abs(+/- 0.60). In this context, a positive value reflects a positive correlation, meaning that as one value rises, so does the other. A negative value reflects a negative correlation, meaning as one value increases, the other decreases.
